# Voice as a Biomarker to Detect Acute Decompensated Heart Failure: Pilot Study for the Analysis of Voice Using Deep Learning Models

**DOI:** 10.1101/2023.09.11.23295393

**Authors:** Jieun Lee, Gwantae Kim, Insung Ham, Kyungdeuk Ko, Soohyung Park, You-Jung Choi, Dong Oh Kang, Jah Yeon Choi, Eun Jin Park, Sunki Lee, Seung Young Roh, Dae-In Lee, Jin Oh Na, Cheol Ung Choi, Jin Won Kim, Seung-Woon Rha, Chang Gyu Park, Eung Ju Kim, Hanseok Ko

**Affiliations:** Division of Cardiology, Department of Internal Medicine, Korea University Guro Hospital, Seoul, Republic of Korea; School of Electrical Engineering, Korea University, Seoul, Republic of Korea

**Keywords:** Voice, Acute Decompensated Heart Failure, Biomarker, Deep Learning Models

## Abstract

**Background:** Acute decompensated heart failure (ADHF) is a systemic congestion state requiring timely management. Admission for ADHF is closely related to the readmission and post-discharge mortality in patients, which makes it imperative to detect ADHF in its early stage.

**Methods:** Patients with ADHF needed admission were eligible for enrollment, and those with respiratory infection, sepsis, lung/vocal cord disease, acute coronary syndrome, or serum creatinine>3mg/dL were excluded. A total of 112 patients were enrolled between July, 2020 and December, 2022. Voice was recorded two times: at admission for ADHF, and at discharge. Patients were asked to phonate five Korean vowels (‘a/e/i/o/u’) for 3 seconds each, and then to repeat the sentence ‘daehan minkook manse’ five times. Low-level audio features were extracted for classification. Then, Mel-Spectrogram was extracted from waveform and used as input features of the deep learning-based classification models. Two kinds of the deep learning-based classification models, convolutional neural networks and Transformer, were adapted for the further analysis.

**Results:** For 100 patients in the final analysis, we randomized patients into two mutually exclusive groups: a training group (n=88) and a test group (n=12). In the analysis with low-level audio features, harmonics-to-noise ratio and Shimmer showed classification potential. Then, deep learning models were trained to classify whether certain voice belongs to ADHF state or recovered state. We treated it as a binary classification task, and the best performing model achieved a classification accuracy of 85.11% with DenseNet201. The classification accuracy was improved as 92.76% with ViT-16-large after inputting additional classic features of heart failure. With adding the low-level audio features in a training process, classification task accuracy was improved in DenseNet201 for about 2%.

**Conclusions:** Our results proposed the clinical possibility of voice as a useful and noninvasive biomarker to detect ADHF in its early stage.

## Introduction

Heart failure (HF) is a clinical syndrome with symptoms and signs due to impairment of heart function and blood supply to the body^1^. More than 64 million people around the world are suffering from HF^2^, and it is estimated to affect more than 15% in those ≥80 years old^3, 4^. HF accounts for the most common cause of hospital admissions in adults above 65 years^5^, and it remains as one of the most costly and disabling diseases causing repetitive admissions and visits to the emergency room^6^. Acute decompensated heart failure (ADHF) is defined as the pulmonary and systemic congestion resulted from increased filling pressures of the heart^7^, which requires timely management, and its mortality rates is around 20% after discharge^8, 9^. Admission for ADHF is closely related to the readmission and post-discharge mortality in HF patients, which makes it crucial to predict the signs of ADHF.

There are several ways to monitor patients with HF via monitoring pulmonary and systemic congestion state: 1) serum biomarkers, 2) imaging techniques. Measurement of serum biomarkers such as N-terminal pro-B-type natriuretic peptide (NT-proBNP) or B-type natriuretic peptide are recommended in the current European and American guidelines ^1, 10^. Soluble suppression of tumorigenicity 2 (sST2), a relatively novel biomarker, is suggested to represent prognosis of HF because this marker is associated with inflammation and fibrosis, which are the important characteristics of HF^11^. Imaging techniques such as chest x-ray and computed tomography (CT) are also important tools providing evidences of HF including pulmonary congestion, pleural effusion and cardiomegaly. However, both biomarkers and imaging techniques are difficult to be repeated whenever needed because of their costs, discomfort of blood sampling and inconvenience of visiting the hospital to undergo such tests. Imaging techniques such as CT can quantify the severity of pulmonary congestion, but because of radiation exposure, it cannot be repeated frequently. As a result, a noninvasive, safe, relatively inexpensive, and accessible test is desperately needed for patients with HF.

Body fluid overload in ADHF can result in change of voice characteristics by causing edema of lung, larynx, and vocal cord. It was observed that phonation threshold pressure, an indicator of pulmonary drive for phonation, was increased in systemic dehydration caused by administering a single 60-mg dose of furosemide in healthy adults^12^. Gaining body weight in patients with HF is one of the indicators of ADHF, but the amount of congestion to cause change in body weight can be a lot more than the amount of pulmonary congestion to bring changes in voice, so monitoring voice to detect congestion state could be a more sensitive and useful indicator than monitoring body weight of patients with HF. Voice is a noninvasive, and easy-to-get biomarker which can be easily monitored even in telemedicine settings to detect pulmonary congestion state in patients with HF, but there are few studies in this area. One study quantified the characteristics of voice in young autistic children^13^. There are few studies investigated voice in cardiovascular diseases. One study identified 2 voice characteristics associated with coronary artery disease^14^, and another study was a pilot study of 10 patients with ADHF investigating several voice measures according to HF symptoms^15^. There is a retrospective, observational study investigating the association of the voice signal characteristics to the clinical outcomes in patients with HF^16^. However, these studies are either comprised of small study population or based on a retrospective design.

We hypothesized that there would be certain differences of voice characteristics in patients with ADHF when admitted to the hospital and after being recovered from ADHF. The objective of our study was to test this hypothesis and furthermore, to find out the potential of voice as a useful and noninvasive biomarker to detect ADHF in its early stage.

## Method

### Study Design and population

This was a prospective study conducted in a tertiary hospital, Korea University Guro Hospital, in Seoul, Korea. Patients with ADHF needed admission were eligible for enrollment, and those with concurrent respiratory infection, sepsis, lung or vocal cord diseases, acute coronary syndrome, coronavirus disease (COVID-19), or serum creatinine >3mg/dL were excluded. Patients who could not speak Korean were also excluded. Patients who were not able to recover from ADHF were dropped out in the final analysis. A total of 112 patients were enrolled between July 1, 2020 and December 31, 2022. This study was approved by the institutional review board of Korea University Guro Hospital (N: 2020GR0356), and the patients were asked to sign the written informed consent.

Voice was recorded total two times: at admission for ADHF, and at discharge after recovered from ADHF. Voice was recorded in a quite environment, with Sony PCM-A10 recorder in a sampling frequency of 48kHz and scale of 24 bit. Patients were asked to phonate five fundamental Korean vowels (‘a/e/i/o/u’) for three seconds each, and then to repeat the sentence ‘daehan minkook manse’ five times. Voice was recorded in a sitting position, or in a supine position when patients were not able to remain seated because of their medical conditions. Routine tests for patients with HF were also gathered at admission and at discharge: Serum NT-proBNP, sST2, BUN, Creatinine, body weight, Visual Analogue Scale (VAS) for dyspnea and general condition. Patients were asked to select VAS grade based on their subjective symptoms of dyspnea and general condition. VAS grade 0 indicates no dyspnea or best general condition, and patients were asked to select VAS grade of 10 if they felt the grade of dyspnea or their general condition is the worst of they have ever experienced.

### Assessment of the Voice

Total 100 patients were eligible for the final analysis. Patients were randomly divided into 2 mutually exclusive groups: a training group (N=88) and a test group (N=12). We trained the models to classify whether certain voice belongs to ADHF state or a recovered state. We treated it as a binary classification task. First, low-level audio features are extracted and assessed if these features are suitable for classification with statistical analysis. Then, Mel-Spectrogram, which is a time-frequency domain feature, was extracted from time domain waveform. Mel-Spectrogram is used as input features of the deep learning-based classification models. We adapted two kinds of the deep learning-based classification models: convolutional neural networks (CNNs) and Transformer.

CNN style classification models, including ResNet50^17^, ConvNeXt^18^, DenseNet201^19^, ResNeXt^20^, and EfficientNet-b7^21^, refine local features of Mel-Spectrogram to find optimal features for ADHF classification during gradient descent steps. Example structure (DenseNet201) of the CNN style classification models is shown in Figure S1. The typical structure of the CNN style classification model consists of the first convolution layer, CNN stem, and a classification layer. Initially, the first convolution layer adjusts channels of the input features. Secondly, the stem extracts hidden features using CNN layers. For DenseNet201, the stem contains four dense layers and three transition layers. Other CNN style classification models have different CNN structures in the stem. The hidden features are converted into ADHF classification results so that humans can understand them by the classification layer.

Transformer style classification models, such as ViT^22^, find optimal attention maps between cropped patches of the Mel-Spectrogram. The optimized features using attention maps focus on more important input features and fewer meaningless features. The structure of the ViT is described in Figure S2. The input features are first cropped into patches and projected into hidden feature tokens. Each hidden feature token represents one truncated patch. Moreover, one classification token is added to hidden feature tokens to calculate attention maps. The hidden feature tokens are refined by attention maps in the multi-head attention layers. Finally, the refined hidden feature of the classification token is converted into ADHF classification results by the classifying layer.

We trained the deep learning-based classification models with AdamW optimizer^23^ with learning rate=0.00001 and cross-entropy loss. To minimize the effects of unique pronunciation of each patient, we cut the voice file into 1 second, resulting the final data samples as 9,442 samples for a training set and 663 samples for a test set. All experiments for the deep learning models were performed with Pytorch library of Python programming language.

### Statistical Analysis

The voice recordings were processed using a range of algorithms to report 4 different low-level speech features, F0, harmonics-to-noise ratio (HNR), Jitter and Shimmer. The paired Student t-test was used to test the null hypothesis that the mean of the low-level speech features at admission and at discharge are equal. In addition, the distributions of change of low-level speech features from admission to discharge were presented to assess usefulness for classification. Furthermore, the linear regression analysis between the selected biomarkers and low-level speech features were performed to investigate linear relationship and usefulness of the features for voice classification.

Since the features extracted by deep learning cannot be summarized into independent statistical variables, the 2-dimensional t-distributed stochastic neighbor embedding (t-SNE) of the hidden features and classification probability were calculated for each training and test sample. Scipy library of Python programming language was used for the statistical analysis. Since the training group and the test group are unpaired, the Welch t-test was used to test the null hypothesis that mean of each characteristic between the training and the test group is equal.

## Results

### Baseline characteristics

Baseline characteristics of a training group (N=88) and a test group (N=12) are presented in Table 1. The training group was slightly older and had higher serum BUN levels compared to the test group. However, there were no significant differences between the two groups in terms of sex, left ventricular ejection fraction (LVEF), history of diabetes mellitus or hypertension, and serum NT-proBNP at admission and at discharge.

**Table 1.**
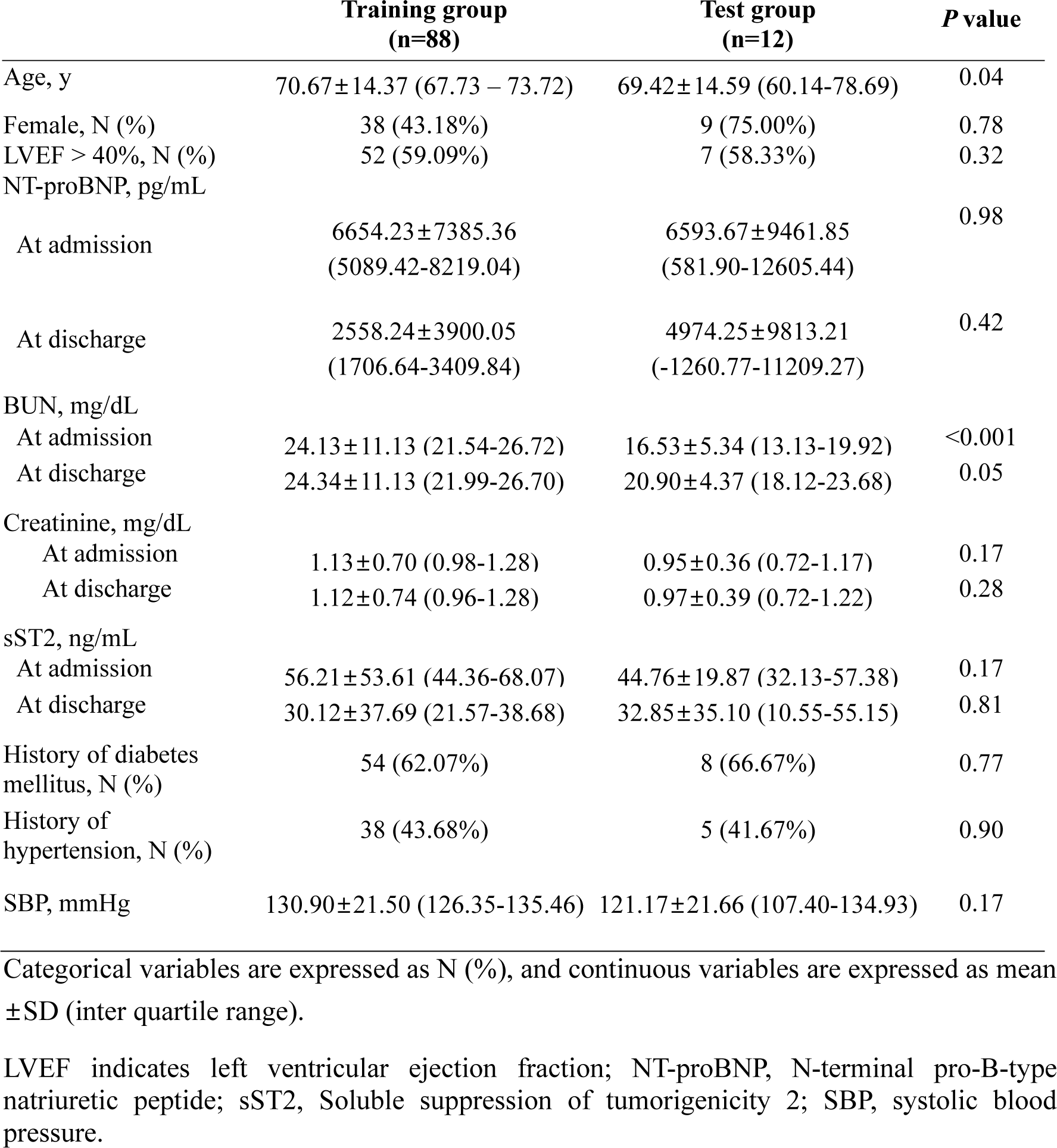
Baseline Characteristics of ADHF Patients.

### Voice analysis

Voice characteristics at admission (‘wet’) and at discharge (‘dry’) were analyzed with low-level audio features to identify certain characteristics of voice that can detect ADHF. Low-level audio features are consisted of F0, HNR, Jitter, and Shimmer. F0 is a frequency, Jitter means consistency of period, and Shimmer reflects consistency of amplitude of voice oscillation. Jitter or Shimmer increases when irregularity of period or amplitude of oscillation increases. HNR measures the ratio between periodic and non-periodic components of a speech sound. Analysis of low-level audio features for ‘daehan minkook manse’ are presented as a box plot in Figure 1. We analyzed the values of low-level audio features in ‘wet’ and ‘dry’ states. Low-level audio features for other pronunciations are shown in Figure S3. There were statistically distinguishable differences in HNR and Shimmer between ‘wet’ and ‘dry’ states for all pronunciation. For ‘u’, Jitter also showed statistically significant differences between ‘wet’ and ‘dry’ states.

**Figure 1.**
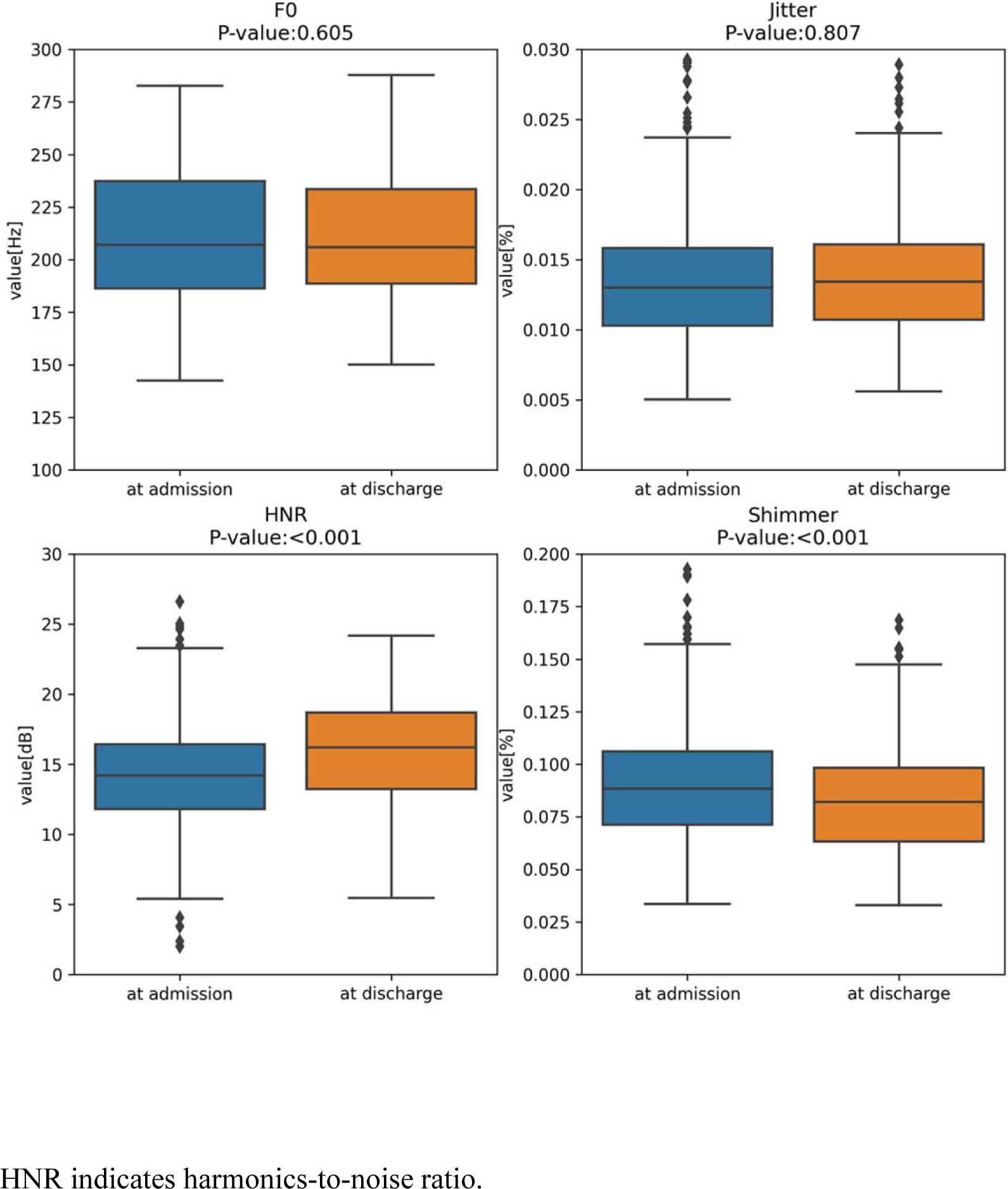
Box Plots of Low-Level Audio Features of ‘daehan minkook manse’.

We further analyzed the change of values of low-level audio features in ‘wet’ and ‘dry’ states. The distribution plots of low-level audio features for ‘daehan minkook manse’ are presented in Figure 2. Since ‘daehan minkook manse’ was recorded five times each at admission and at discharge, the values between different ‘wet’ states and different ‘dry’ states were also analyzed. The distribution of ‘wet-dry’ should be separated from the distribution of ‘wet-wet’ and ‘dry-dry’ states to be a useful feature for voice classification. When we compared the distributions of ‘wet-dry’, ‘wet-wet’, and ‘dry-dry’ states, distributions of the ‘wet-dry’ were not clearly distinct from ‘wet-wet’ and ‘dry-dry’ for Jitter and Shimmer. The average and the distributions of the ‘wet-dry’ were slightly separated from ‘wet-wet’ and ‘dry-dry’ for F0 and HNR, but not enough for classification. The distribution plots of low-level audio features for other pronunciations are presented in Figure S4. Since ‘a/e/i/o/u’ were recorded once each at admission and at discharge, the values between different ‘wet’ states and different ‘dry’ states could not be analyzed. Therefore, if the average value is far from 0 and the distribution plot is not wide, that feature could be useful for classification. As shown in Figure S4, all selected low-level audio features were not suitable for voice classification.

**Figure 2.**
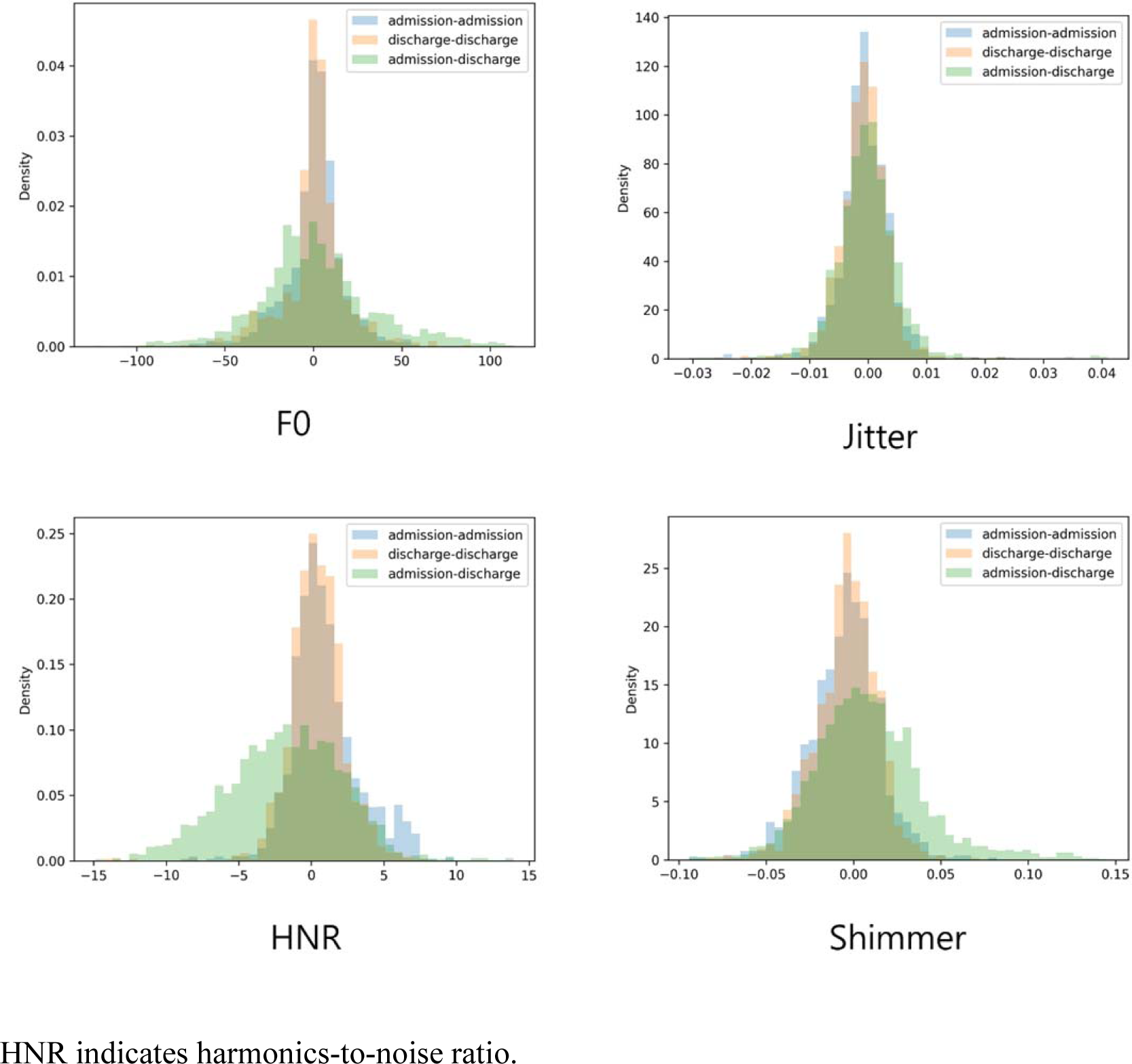
Distribution Plots of Low-Level Audio Features of ‘daehan minkook manse’.

We further conducted the linear regression with absolute values of ‘wet’ and ‘dry’ states by inputting audio features and classic serum biomarkers of HF (NT-proBNP and sST2) for ‘deahan minkook manse’ (Figure 3). Because of many outliers, it can be inferred that low-level audio features and serum biomarkers do not have linear associations. Also, distributions of ‘wet’ and ‘dry’ states were almost overlapped. Linear regression plots of other pronunciations are presented in Supplemental Figure 5.

**Figure 3.**
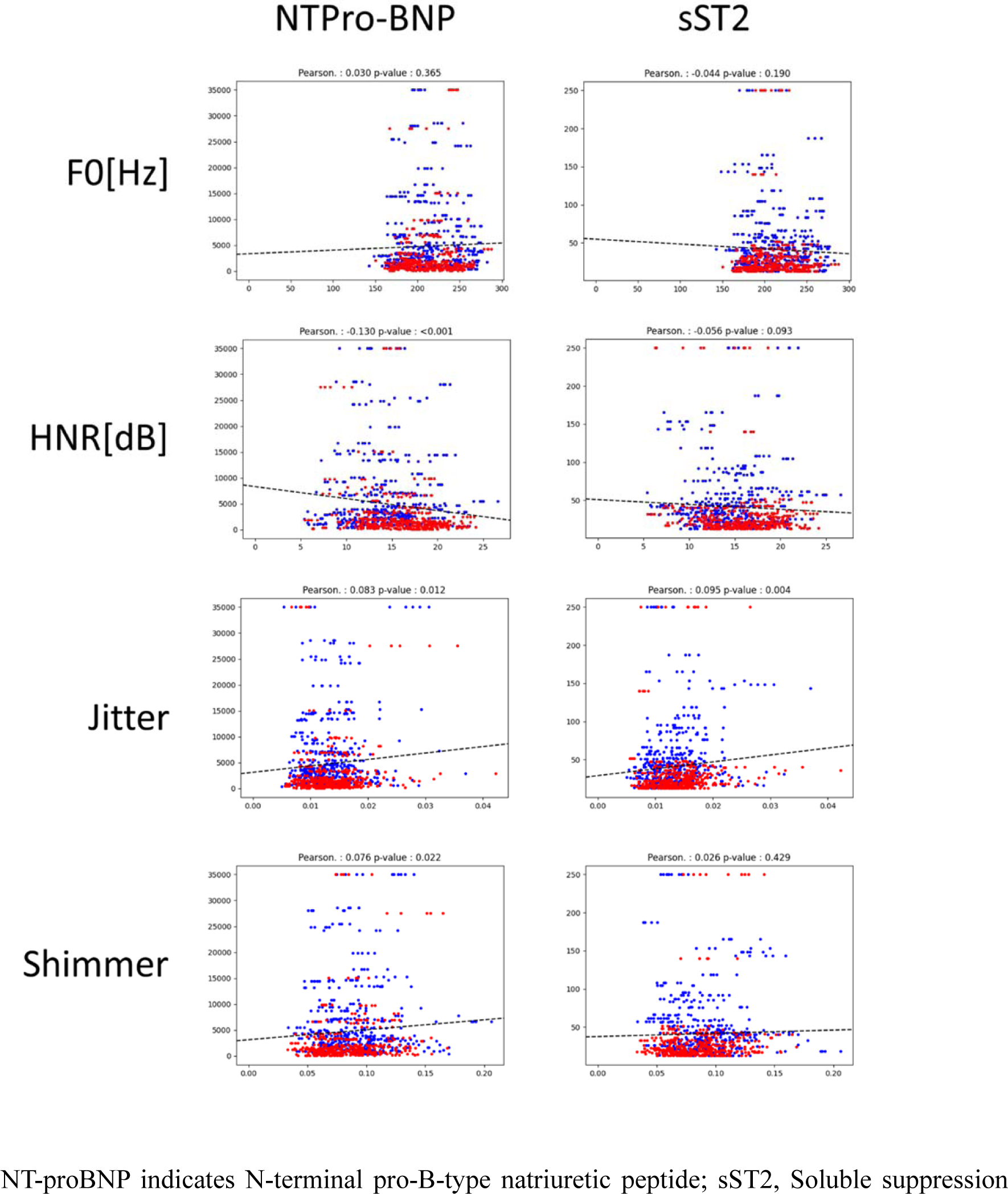
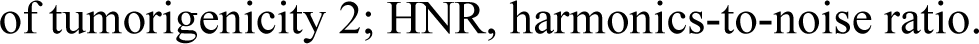
Linear Regression of Low-Level Audio Features and Serum Biomarkers for ‘daehan minkook manse’.

Since analysis with low-level voice features were not significantly distinct between ‘wet’ and ‘dry’ states, we further analyzed voice with Mel-Spectrogram, CNNs, and deep learning models. Mel-Spectrogram of the training set for each word are shown in Figure S6. Time-frequency feature of Mel-Spectrogram were as follows: N-FFT = 1,024, hop_length = 256, mel_bin = 128. The t-SNE plots of all training and test samples are shown in Figure 4. The 256-dimensional features are projected into 2-dimension for visualization. The hidden features at admission and at discharge were not clearly distinct from each other. The representative Mel-Spectrograms with the highest and the lowest classification scores for ‘dry’ states are presented in Figure S7. The Mel-Spectrogram at admission is noisy and the Mel-Spectrogram at discharge is relatively clean with a classification score of 0.999.

**Figure 4.**
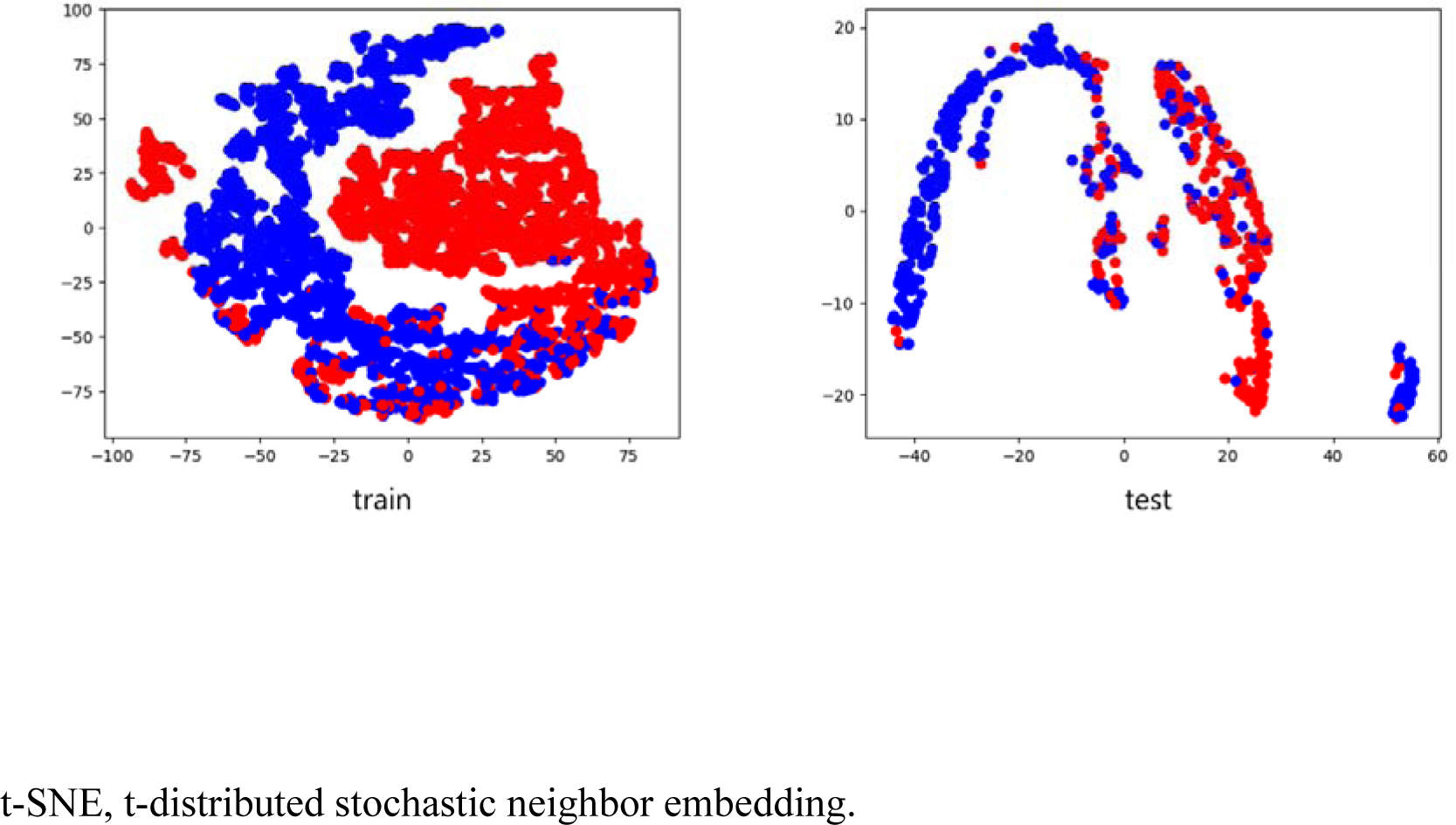
t-SNE Plots of the Last Hidden Features in Trained DenseNet201 for ‘daehan minkook manse’. Red: at admission. Blue: at discharge.

After classifying voice by CNNs, we trained the deep learning models for 5 times with 1-second voice files of ‘daehan minkook manse’, and then a binary classification task was given to each model to classify whether certain voice belongs to ADHF state or recovered state (Table 2). The best performing model achieved an average classification accuracy of 85.11% with DenseNet201. Average classification accuracy was improved to 92.76% with ViT-16-large after inputting additional classic features of HF including NT-proBNP, sST2, VAS of dyspnea & general condition, and body weight. For all pronunciation including ‘a/e/i/o/u’, DenseNet201 achieved the best classification accuracy of 82.38%. DenseNet201 was still the best performing model after inputting additional features of HF with mild improvement to 83.98% of classification accuracy. When we combined DenseNet201 with ViT-16-large, the classification accuracy was lowered when training only with voice (79.37%), but the classification accuracy of this combination model significantly improved (84.80%) after training further with additional HF features.

**Table 2.**
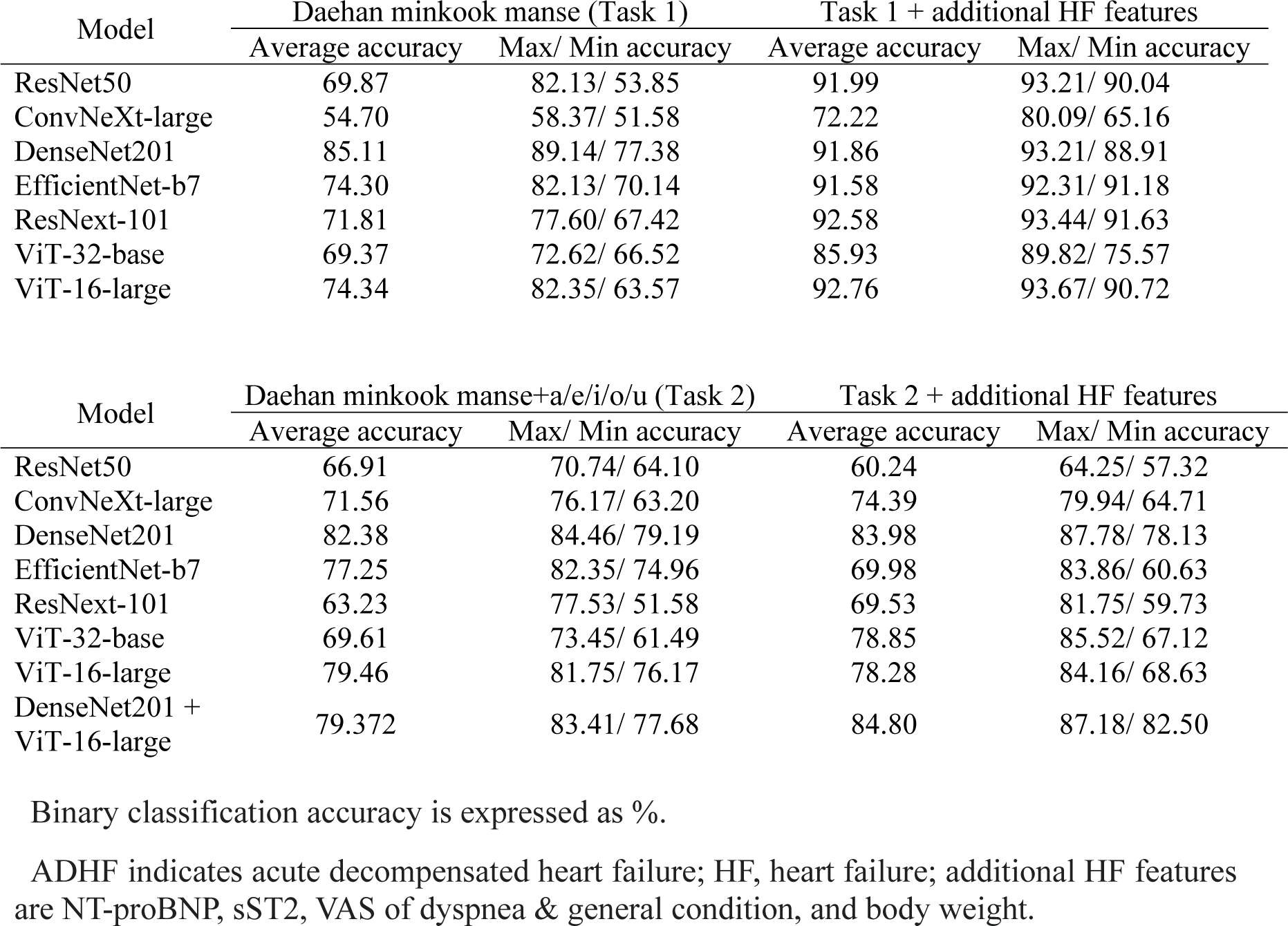
Binary Classification Task Accuracy of Distinguishing Between ADHF and Recovered States with Deep Learning-Based Classification Models.

| Model | Daehan minkook manse (Task 1) |  | Task 1 + additional HF features |  |
| --- | --- | --- | --- | --- |
|  | Average accuracy | Max/ Min accuracy | Average accuracy | Max/ Min accuracy |
| ResNet50 | 69.87 | 82.13/ 53.85 | 91.99 | 93.21/ 90.04 |
| ConvNeXt-large | 54.70 | 58.37/ 51.58 | 72.22 | 80.09/ 65.16 |
| DenseNet201 | 85.11 | 89.14/ 77.38 | 91.86 | 93.21/ 88.91 |
| EfficientNet-b7 | 74.30 | 82.13/ 70.14 | 91.58 | 92.31/ 91.18 |
| ResNext-101 | 71.81 | 77.60/ 67.42 | 92.58 | 93.44/ 91.63 |
| ViT-32-base | 69.37 | 72.62/ 66.52 | 85.93 | 89.82/ 75.57 |
| ViT-16-large | 74.34 | 82.35/ 63.57 | 92.76 | 93.67/ 90.72 |

| Model | Daehan minkook manse+a/e/i/o/u (Task 2) |  | Task 2 + additional HF features |  |
| --- | --- | --- | --- | --- |
|  | Average accuracy | Max/ Min accuracy | Average accuracy | Max/ Min accuracy |
| ResNet50 | 66.91 | 70.74/ 64.10 | 60.24 | 64.25/ 57.32 |
| ConvNeXt-large | 71.56 | 76.17/ 63.20 | 74.39 | 79.94/ 64.71 |
| DenseNet201 | 82.38 | 84.46/ 79.19 | 83.98 | 87.78/ 78.13 |
| EfficientNet-b7 | 77.25 | 82.35/ 74.96 | 69.98 | 83.86/ 60.63 |
| ResNext-101 | 63.23 | 77.53/ 51.58 | 69.53 | 81.75/ 59.73 |
| ViT-32-base | 69.61 | 73.45/ 61.49 | 78.85 | 85.52/ 67.12 |
| ViT-16-large | 79.46 | 81.75/ 76.17 | 78.28 | 84.16/ 68.63 |
| DenseNet201 + ViT-16-large | 79.372 | 83.41/ 77.68 | 84.80 | 87.18/ 82.50 |

Binary classification accuracy is expressed as %.
ADHF indicates acute decompensated heart failure; HF, heart failure; additional HF features are NT-proBNP, sST2, VAS of dyspnea & general condition, and body weight.

Additional binary classification tasks were conducted in DenseNet201 and ViT-16-large, the two best performing models in classification accuracy, with inputting low-level audio features (Table 3). With adding the low-level audio features in a training process, classification task accuracy was improved in DenseNet201 for about 2%, whereas classification accuracy was deteriorated for about 2% in ViT-16-large model.

**Table 3.**
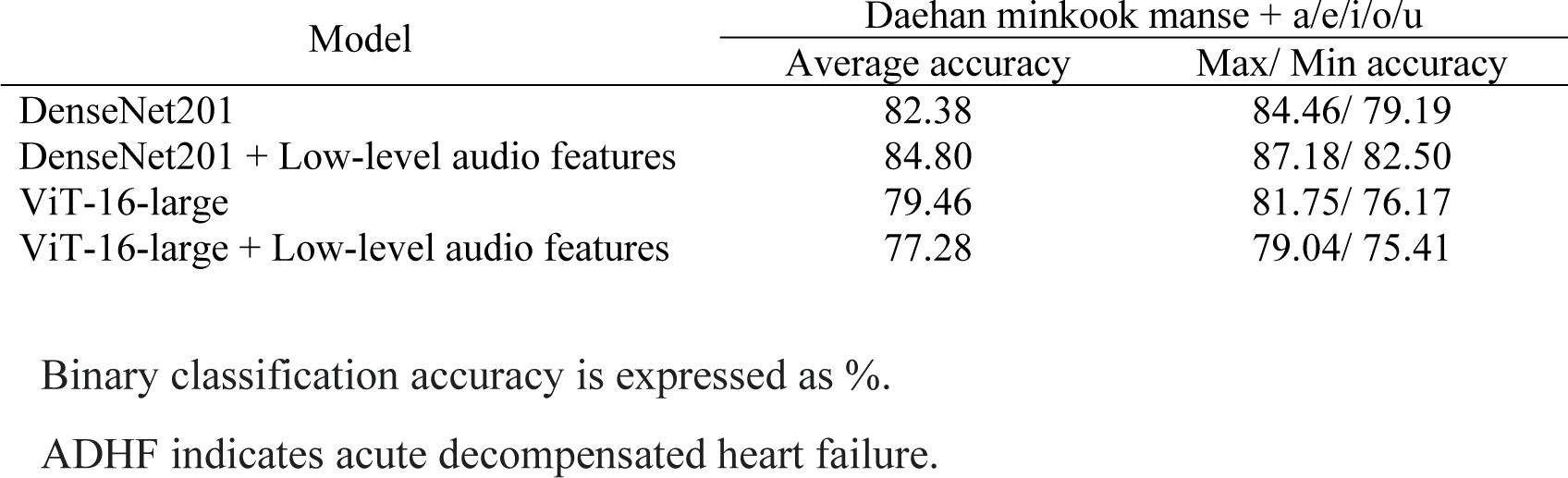
Binary Classification Task Accuracy of Distinguishing Between ADHF and Recovered States Using Deep Learning-Based Classification Models and Low-Level Audio Features.

## Discussion

This is the first study to demonstrate clinical possibility of voice as a biomarker to detect ADHF using deep learning models. We trained the models to classify whether certain voice belongs to ADHF state or a recovered state. Even though low-level audio features showed limited classification potential, further analysis with Mel-Spectrogram and deep learning-based models achieved the classification accuracy over 80%. This classification accuracy was improved to the maximum 92.76% with inputting additional features of HF including NT-proBNP, sST2, VAS of dyspnea & general conditions, and body weight.

There have been a few attempts to investigate the potential of voice as a biomarker for HF. One pilot study analyzing several voice measures in 10 patients with ADHF presented certain tendencies in some features, but many results were not uniform possibly because of small study population^15^. One retrospective study investigated certain voice characteristics associated with adverse outcomes among patients with congestive HF, but the study population was patients who were in chronic conditions, not in ADHF states^16^. Recently, there was a study analyzing voice recordings of 40 patients using a speech processing application^24^. Although it was a prospective study of patients with ADHF, it merely analyzed differences between baseline recording and discharge recordings. In our study, we tried to adapt more realistic approaches. Since the purpose of our study is to find out the possibility of voice as a useful biomarker to detect ADHF in its early stage for timely management before it is too late, we trained the deep learning models with voice of a training group, and tested them to undergo a binary classification task for a test group. The best performing model achieved classification accuracy as high as 85.11%, and the classification accuracy was improved with inputting additional classic features of HF.

Voice is produced as a cooperative result of the lungs, the larynx, and the articulators^25^. Patients with ADHF have pulmonary congestion and systemic edema, which can affect the conditions of the articulatory organs. This swelling of articulatory organs including vocal cord, tongue, lips, and cheeks can bring changes in voice and speech production. Although findings of this study cannot explain a direct mechanism of voice changes in ADHF patients, our study presented a clinical potential of voice as a useful biomarker to detect ADHF by achieving classification accuracy more than 80%. With adding classic features of HF including NT-proBNP, sST2, VAS of dyspnea & general condition, and body weight, the classification accuracy was significantly improved, which could suggest clinical practicality of the selected deep learning models of our study. Although analysis with low-level voice features were not significantly distinct between ‘wet’ and ‘dry’ states, when these low-level features were added in a binary classification task performed by the two best performing models, classification task accuracy was improved in DenseNet201, while it was deteriorated in ViT-16-large model. This could suggest possibility of low-level voice features in a classification process with carefully chosen deep learning models.

This study has several limitations. First, only a selected population of patients who can speak Korean were included in this study. Even though we have tried to minimize the effect of differences in pronunciation by cutting the voice recordings into 1 second, further studies may be needed to verify our findings in the other patient groups who speak different languages. Second, since the voice saying given phrases must be recorded from patients with ADHF, patients in intensive care units were not able to participate in the study because of their intubated states or huge noises from the surrounding machines such as ventilators, continuous renal replacement therapy machines, cardiac monitors, and defibrillators. Third, even though we included relatively large number of patients compared to the previous studies, future studies with larger patient population might bring better classification accuracy. Lastly, because this study was conducted during COVID-19 pandemic and patients with COVID-19 were excluded, a selection bias was inevitable.

## Conclusions

After training deep learning models to classify whether certain voice belongs to ADHF state or recovered state, the best performing model achieved a classification accuracy of 85.11%, and this classification accuracy was improved to higher than 90% with adding classic features of HF. In future study, we plan to conduct the deep learning model structures that address more hidden features in voice for classifying ADHF with more diverse pronunciations and larger patient population. We believe the findings of our study could help patients with HF to be managed earlier before falling into serious ADHF states, furthermore help them to live with better quality of life and have less mortality or morbidity from ADHF.

## Data Availability

The data generated in this study is available from the corresponding author upon reasonable request.

## Nonstandard Abbreviations and Acronyms

ADHF: acute decompensated heart failure
CNNs: convolutional neural networks
COVID-19: coronavirus disease 2019
CT: computed tomography
EF: ejection fraction
HF: heart failure
HNR: harmonics-to-noise ratio
NT-proBNP: N-terminal pro-B-type natriuretic peptide
sST2: Soluble suppression of tumorigenicity 2
t-SNE: t-distributed stochastic neighbor embedding
VAS: Visual Analogue Scale

## Acknowledgments

The authors thank the patients with ADHF for participating in this study.

## Supplemental Material

**Supplemental Figure 1.**
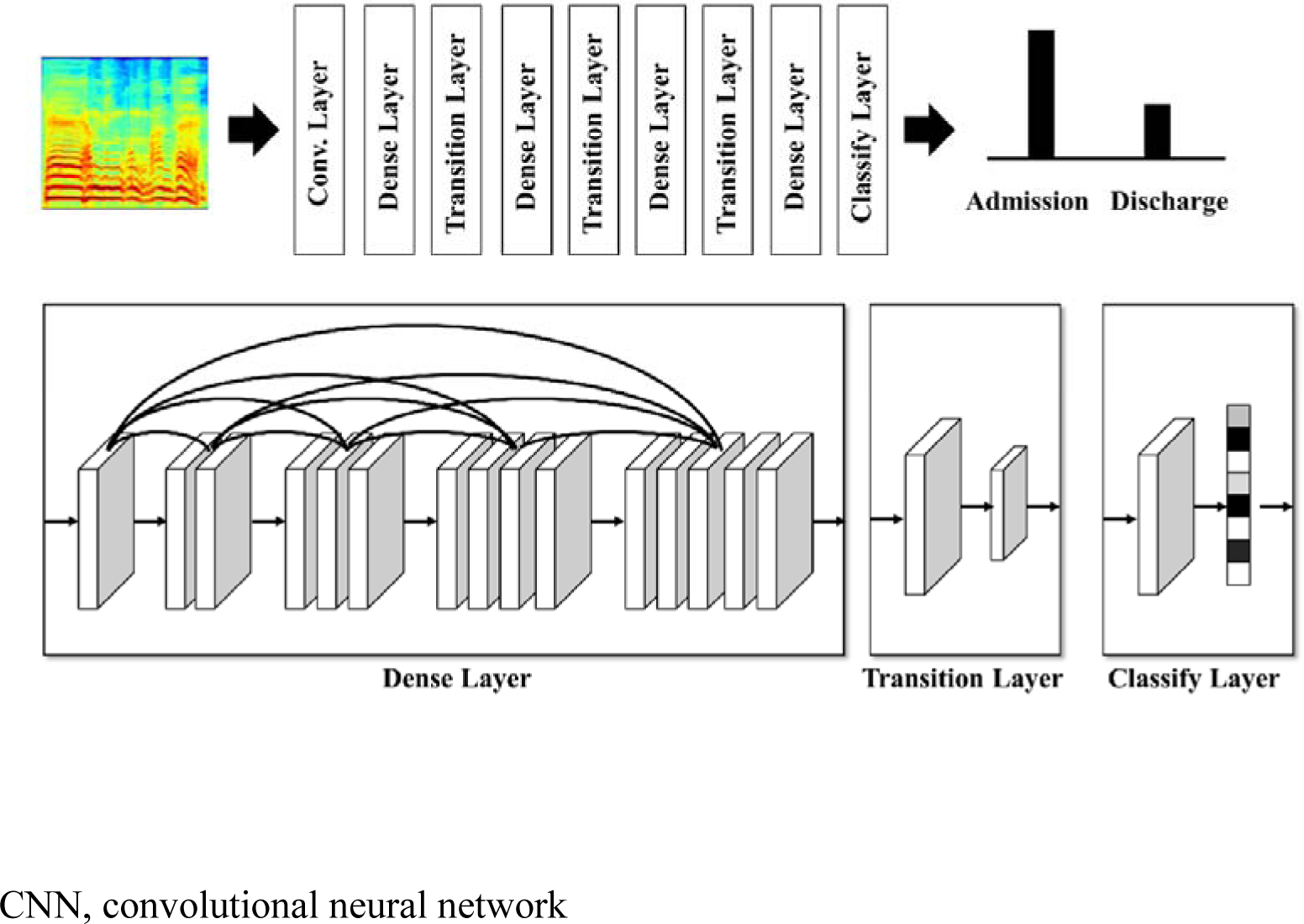
CNN Structure Model (DenseNet201)

**Supplemental Figure 2.**
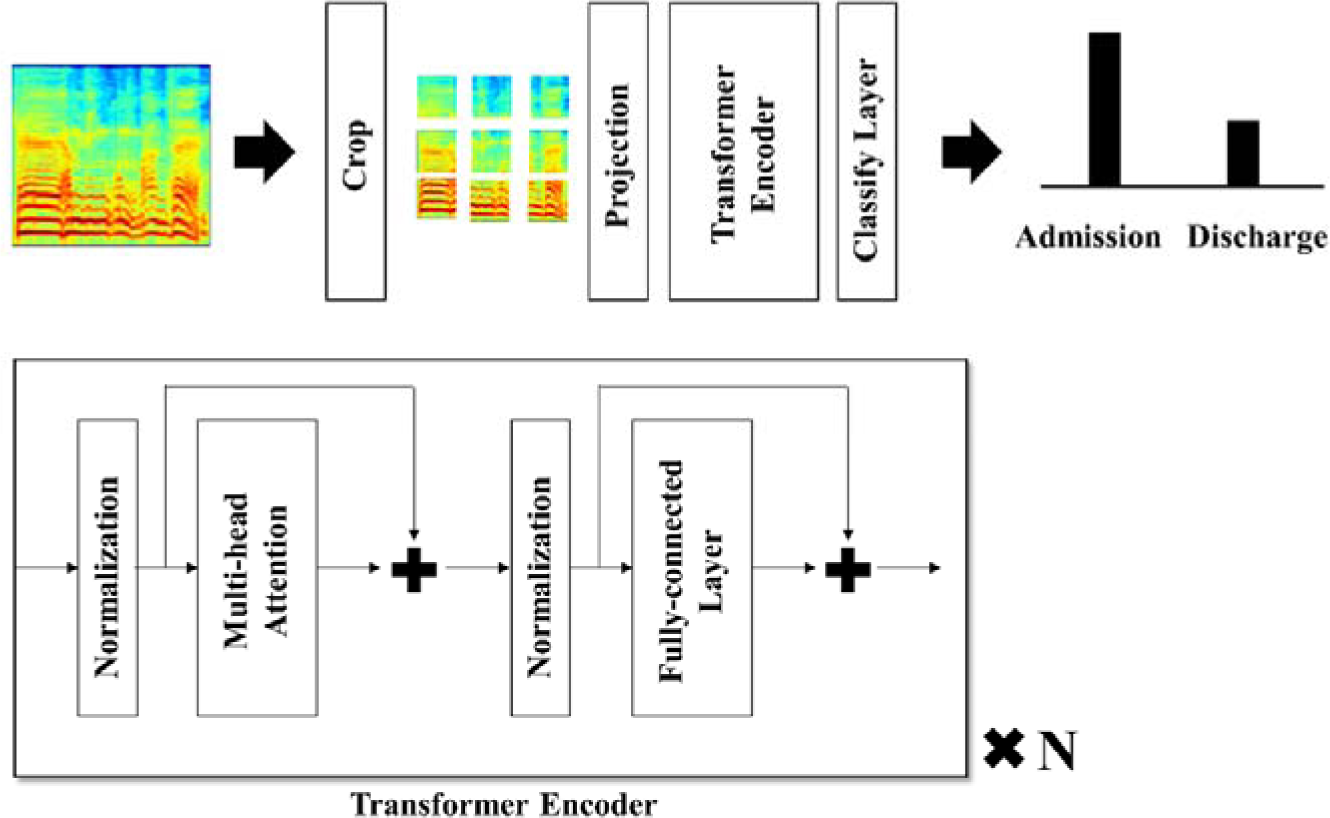
Transformer Style Classification Model (ViT)

**Supplemental Figure 3.**
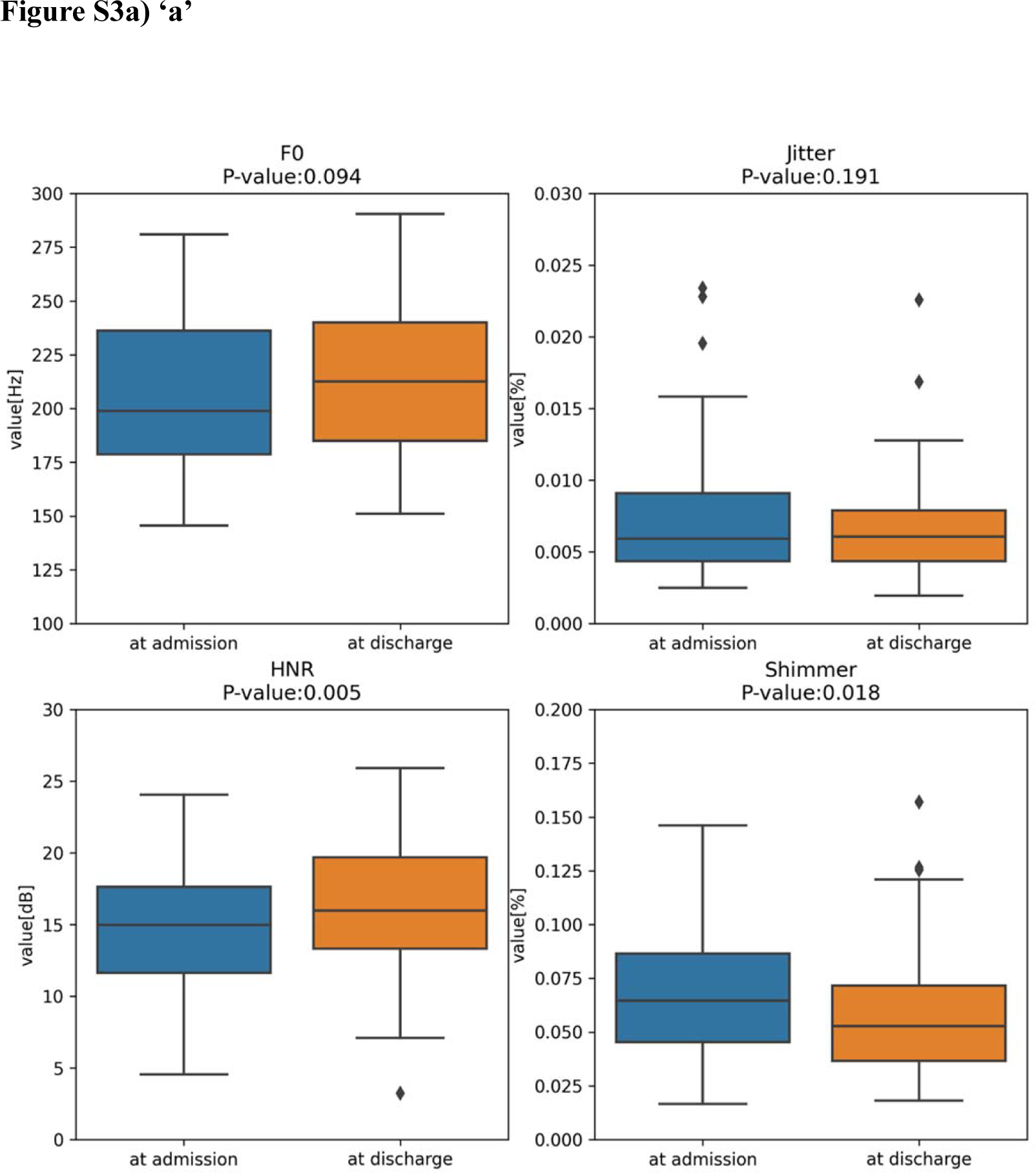
Box Plots of Low-Level Audio Features For ‘a/e/i/o/u’. HNR, harmonics-to-noise ratio.

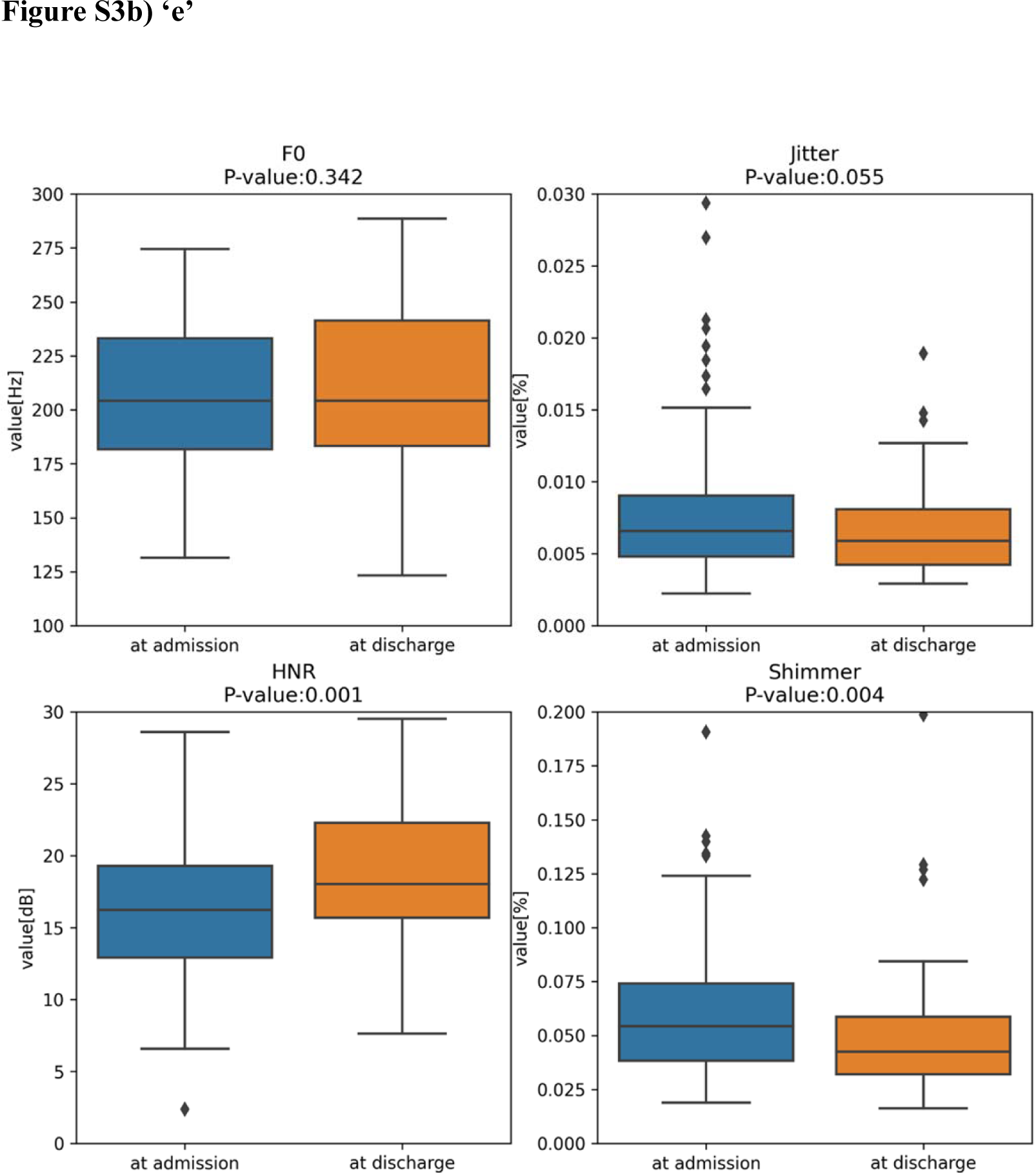
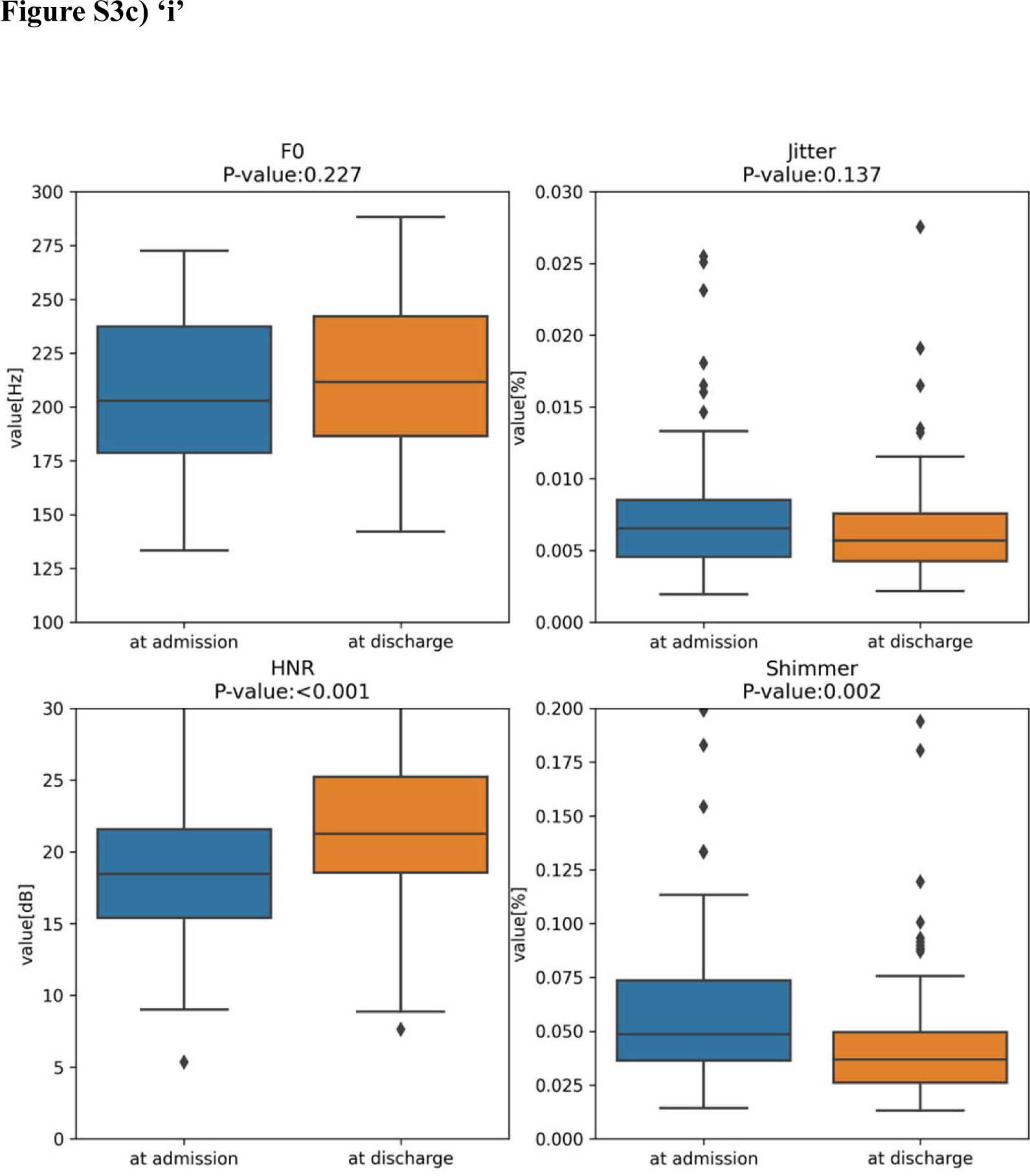
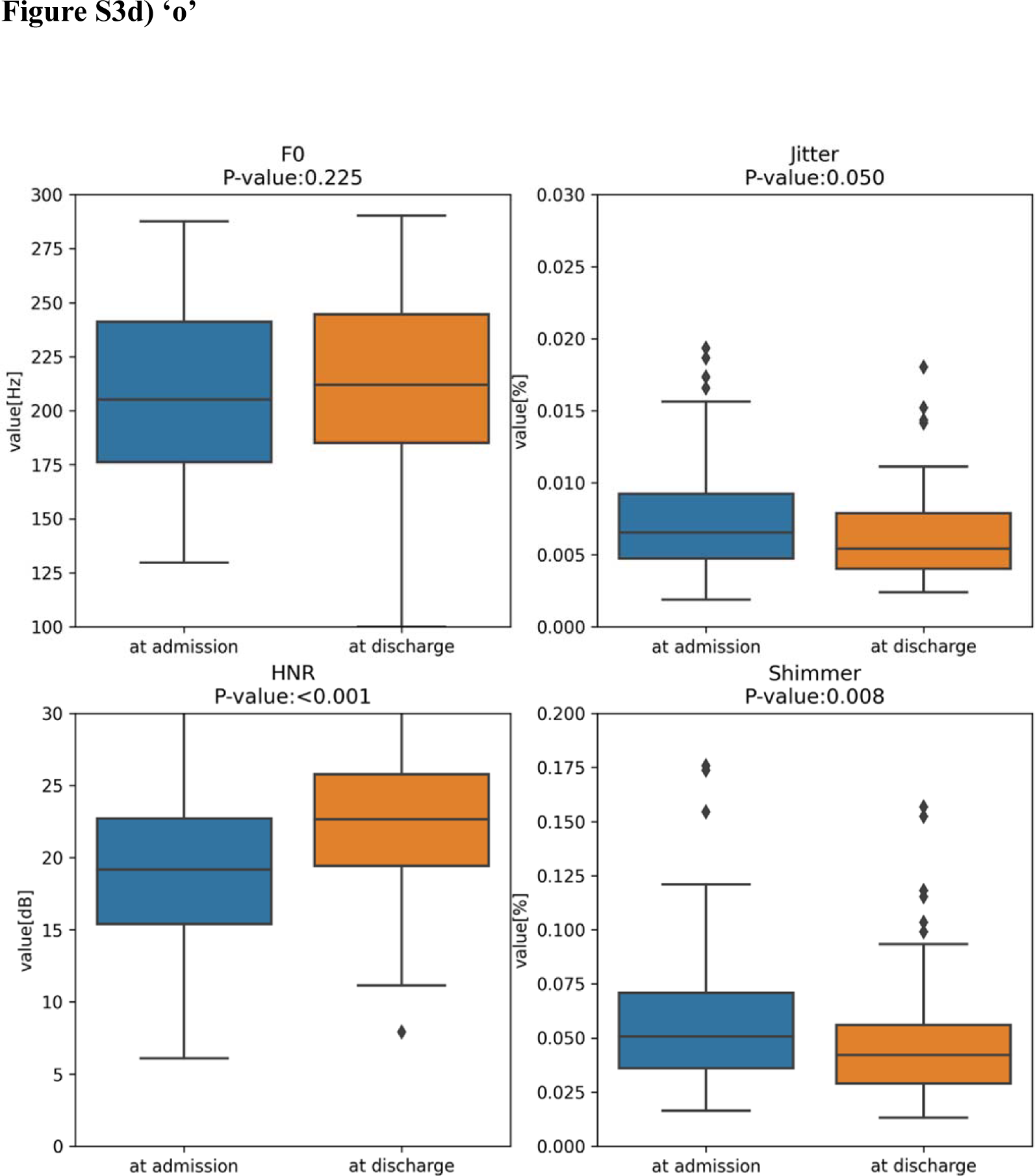
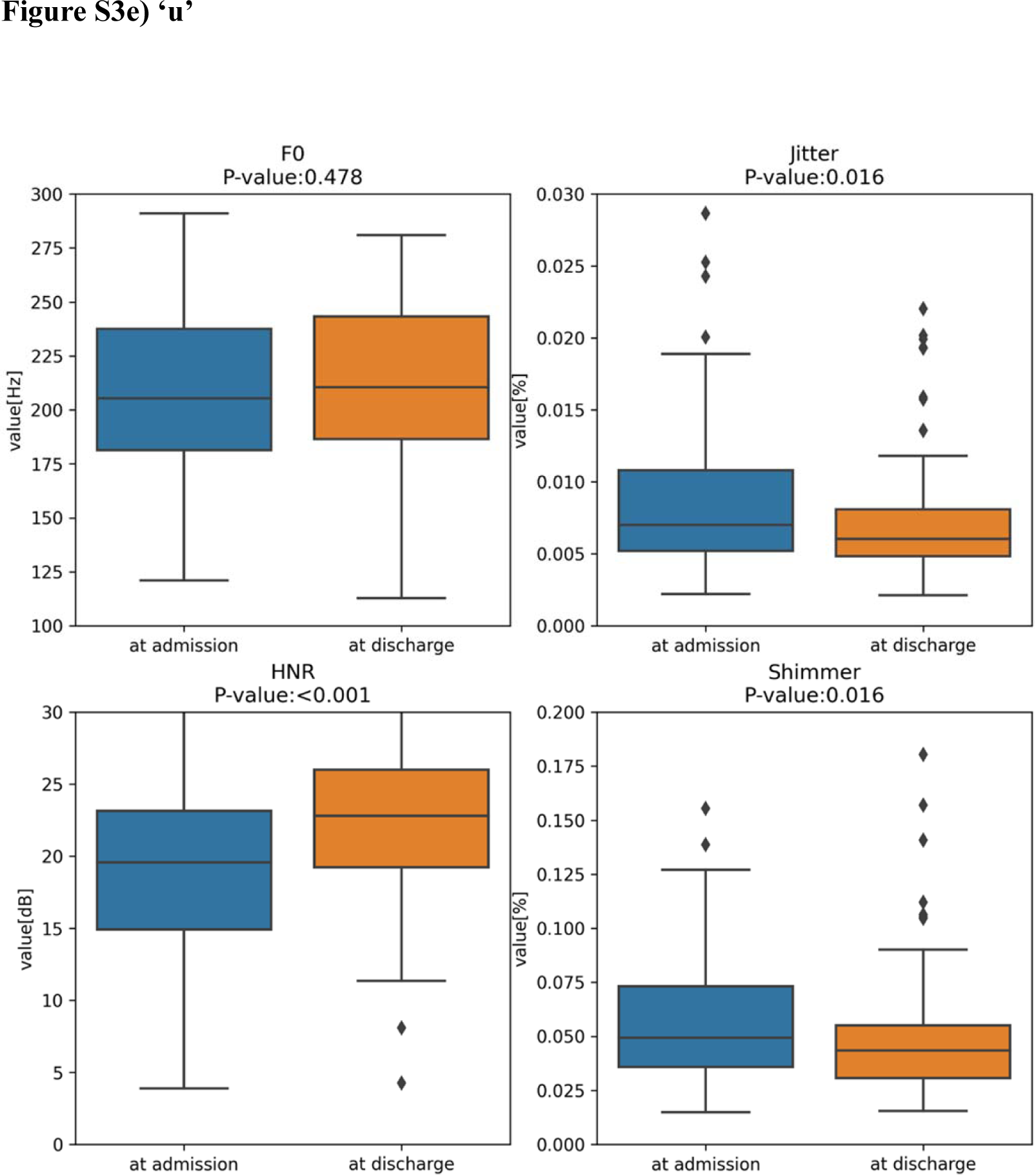

**Supplemental Figure 4.**
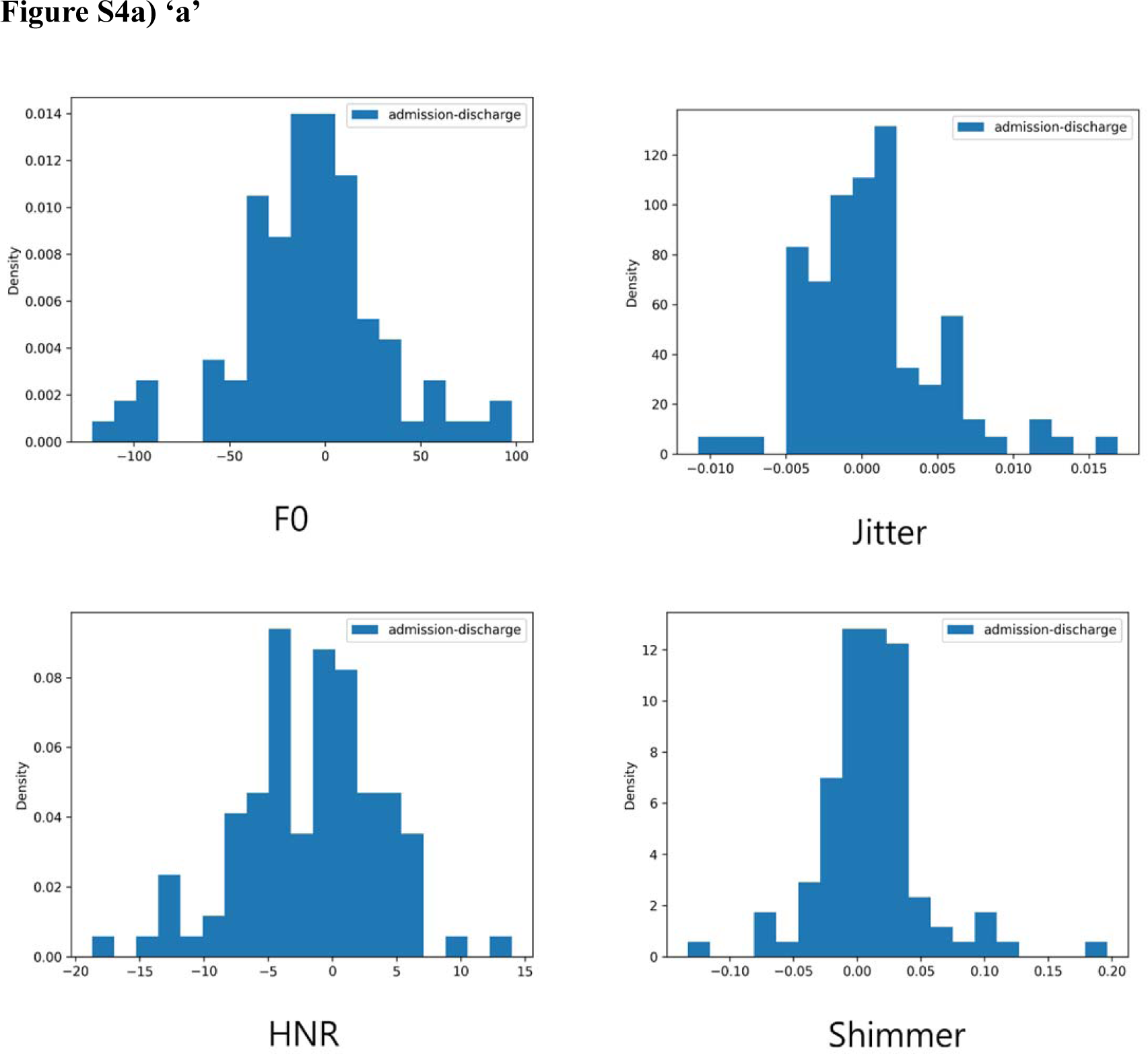
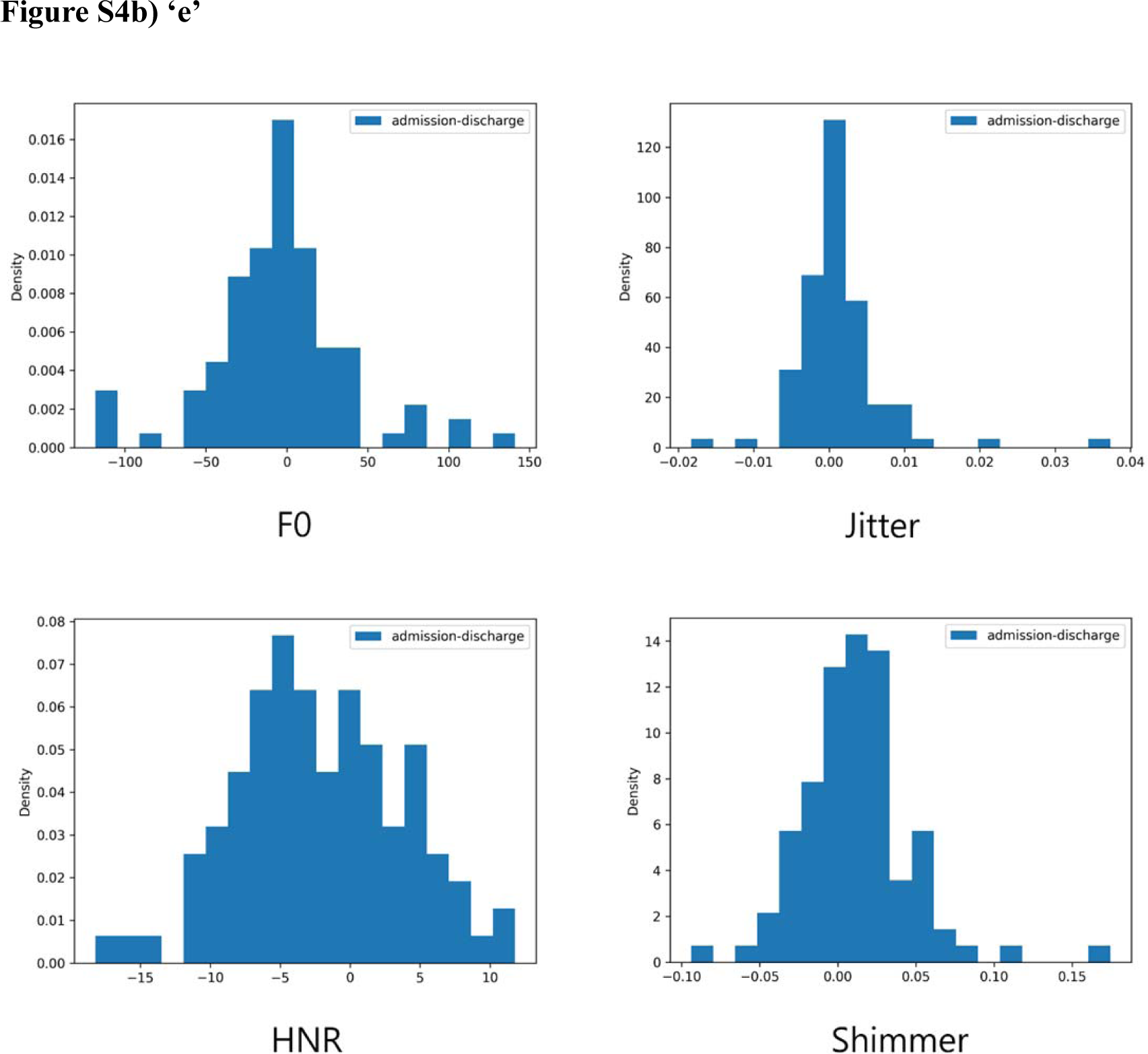
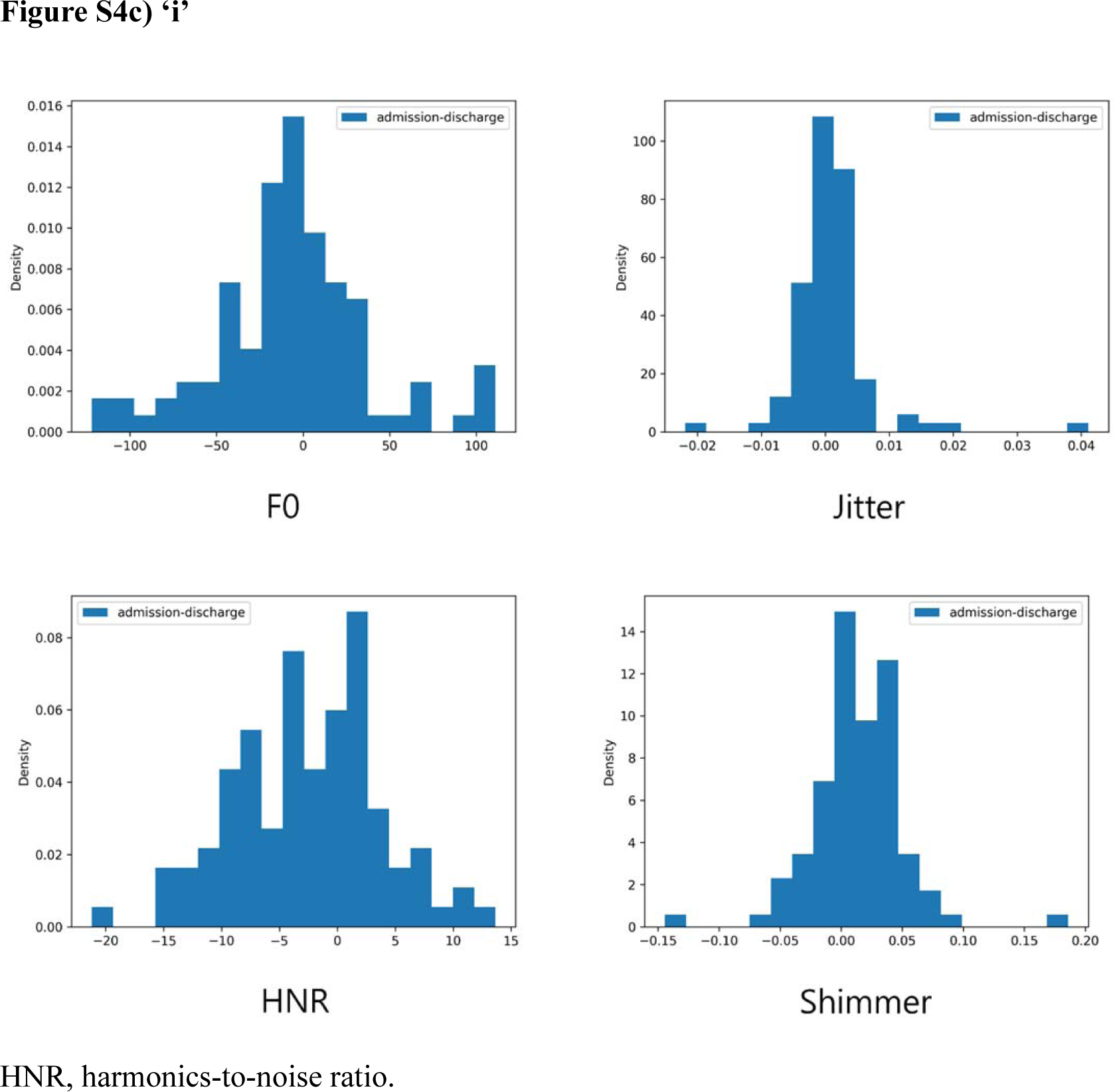
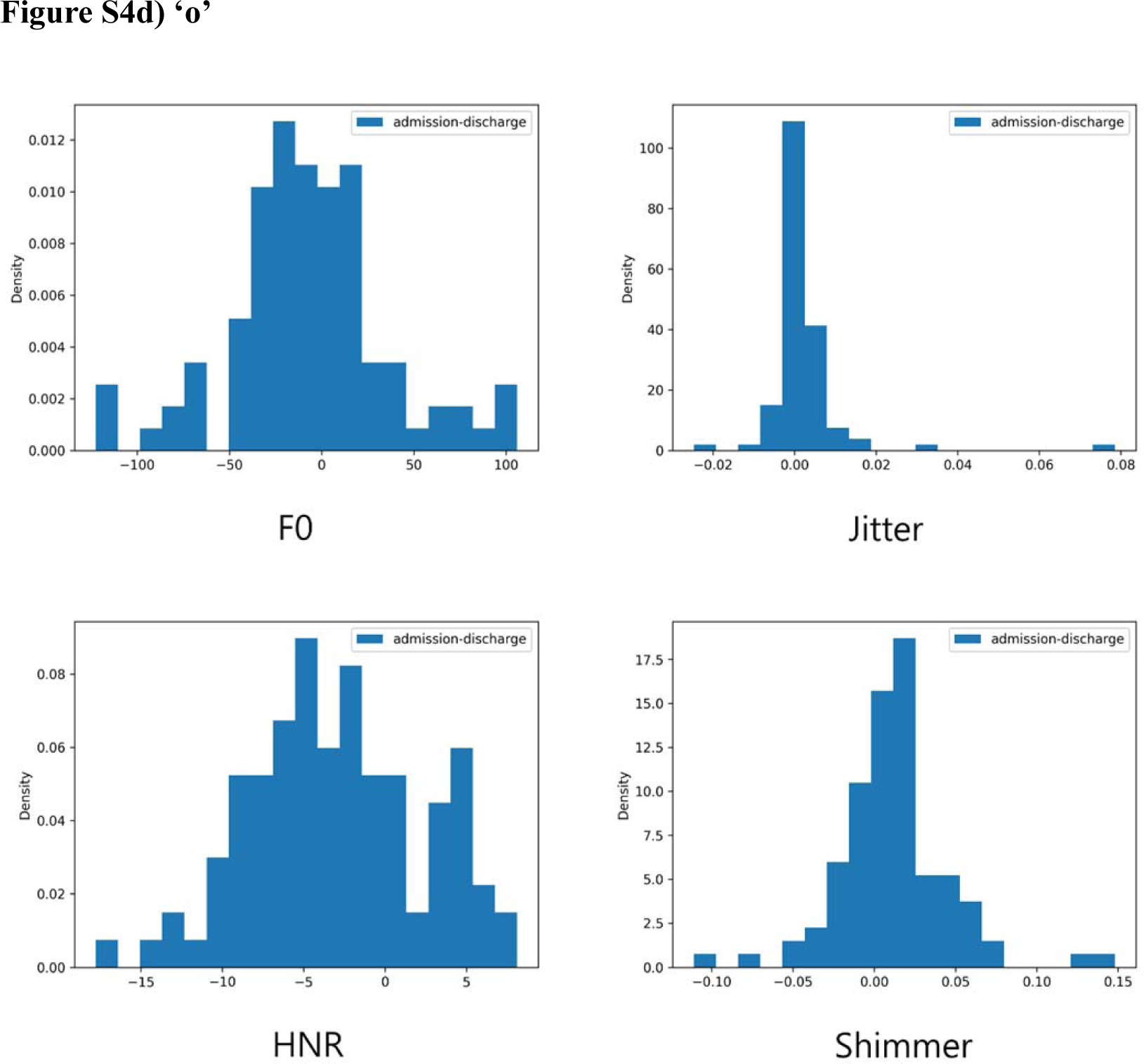
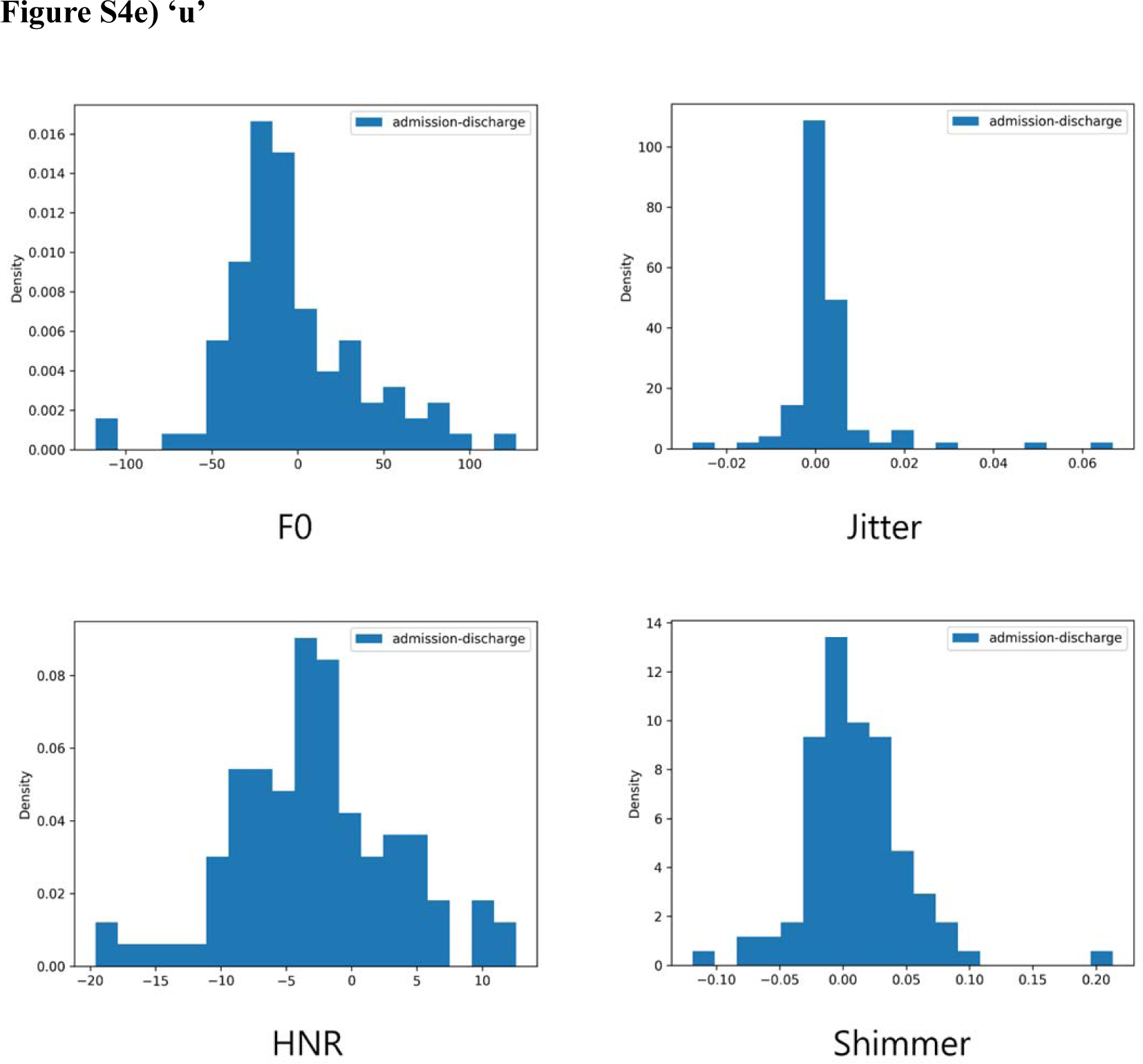
Distribution plot of Low-Level Audio Features For ‘a/e/i/o/u’. HNR, harmonics-to-noise ratio.

**Supplemental Figure 5.**
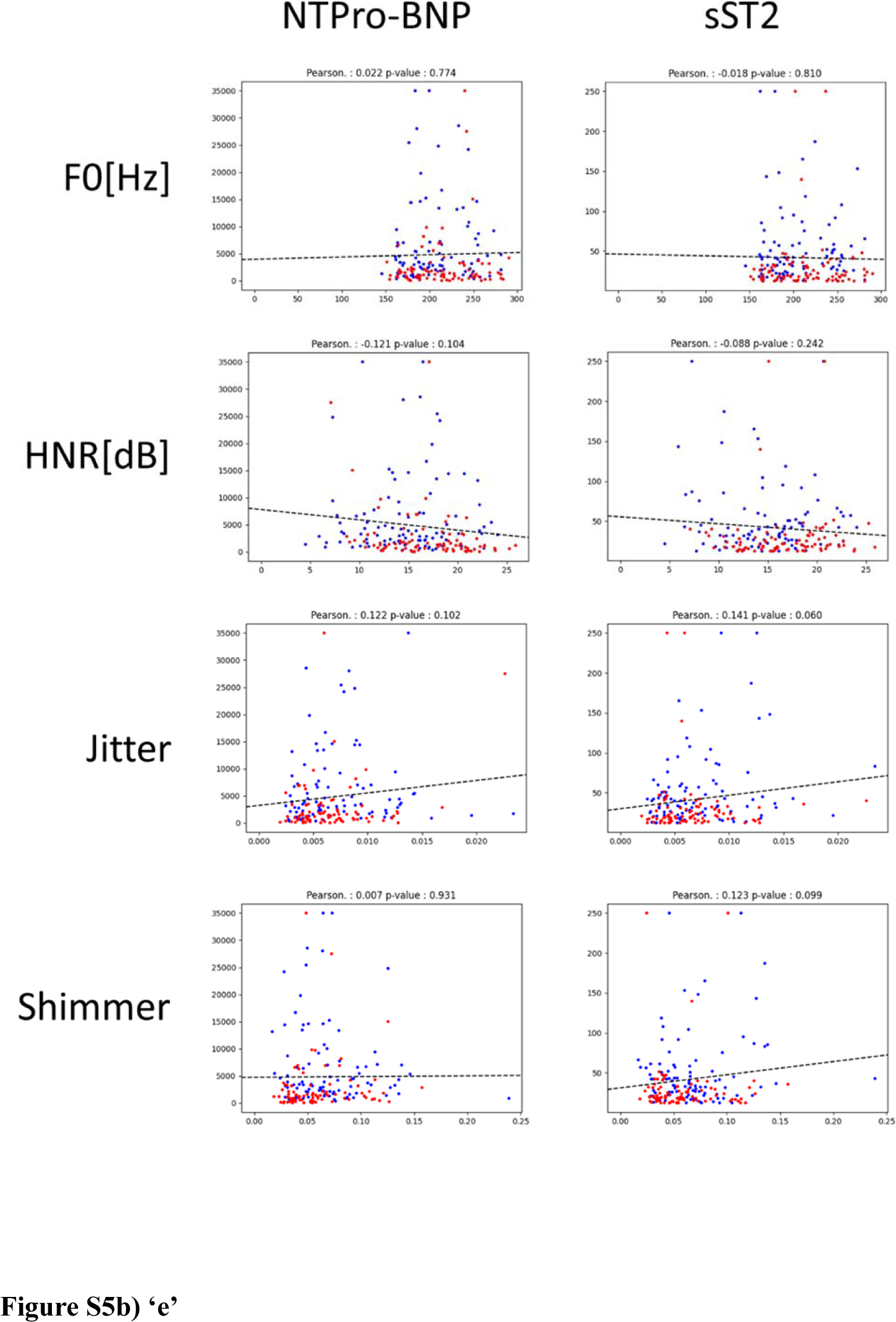
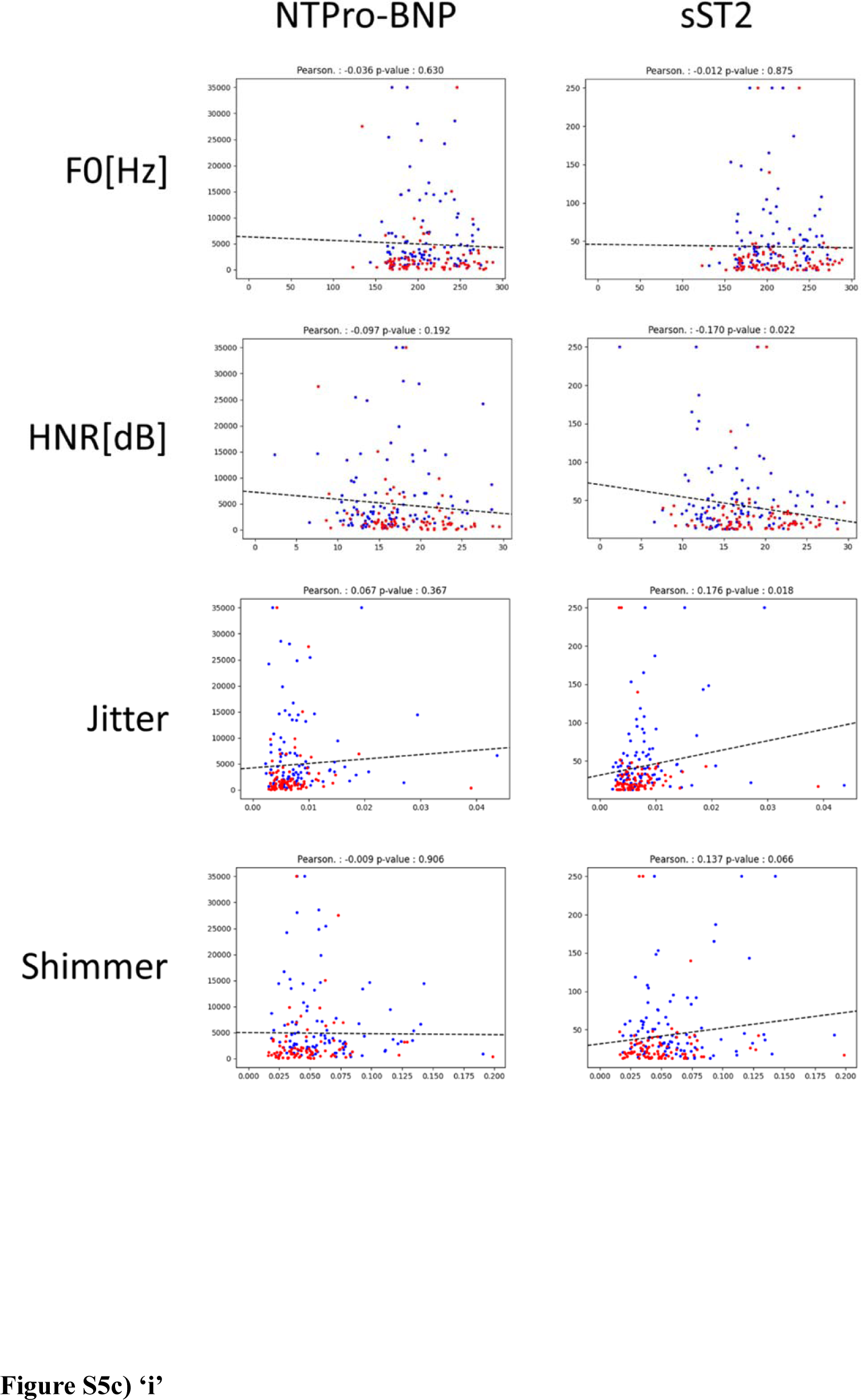
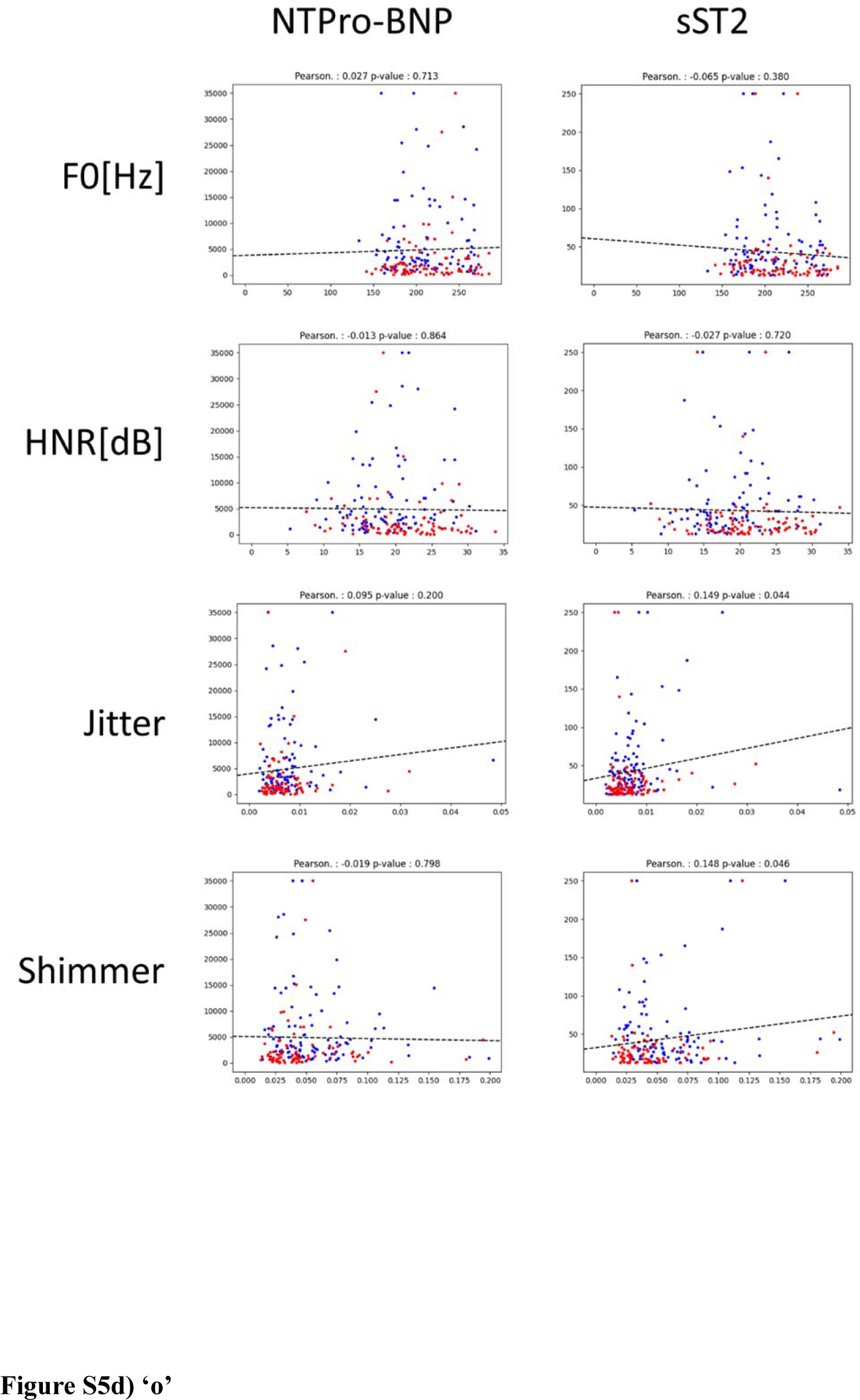
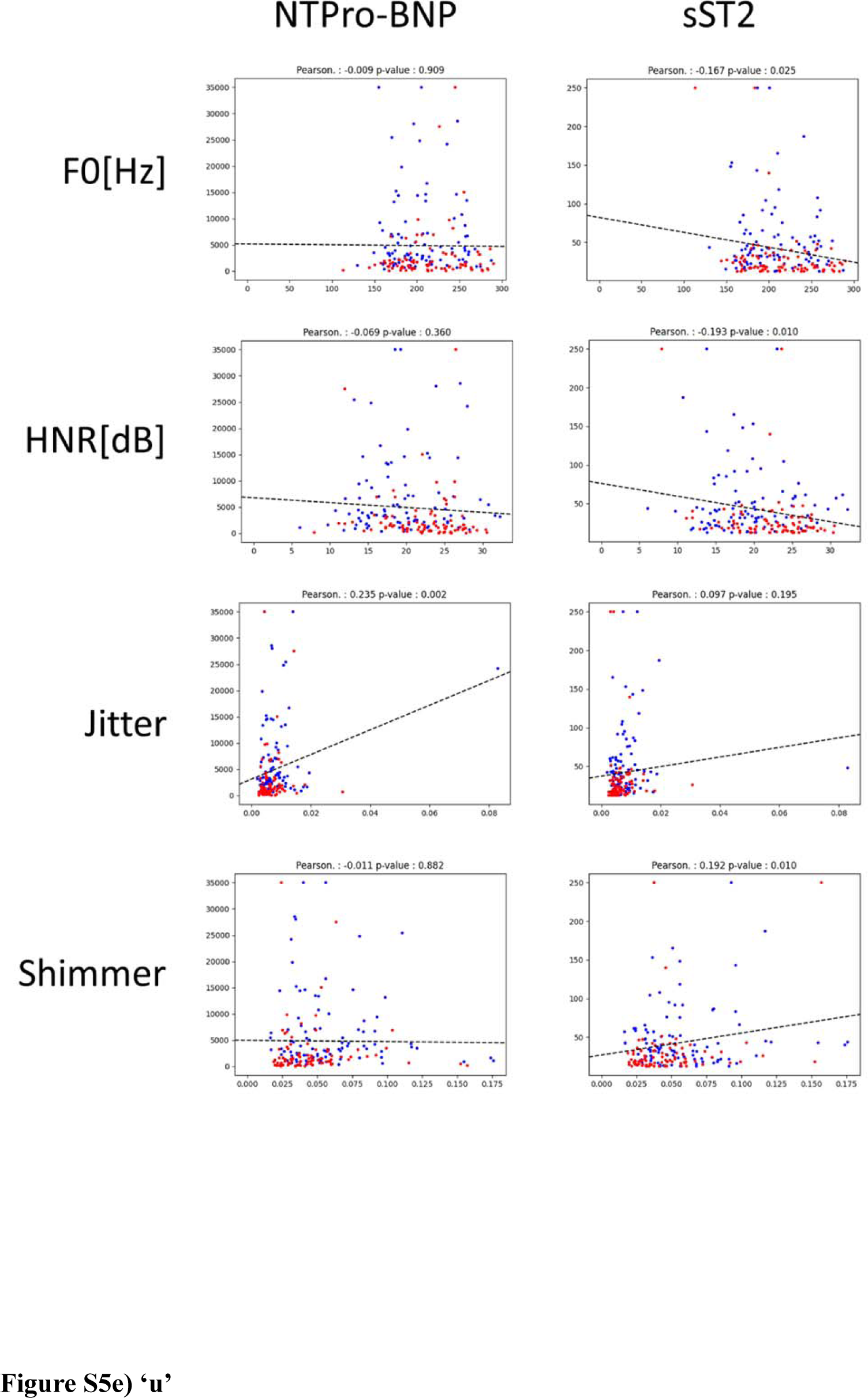
Linear Regression of Low-Level Audio Features and Serum Biomarkers for ‘a/e/i/o/u’. HNR, harmonics-to-noise ratio. **Figure S5a) ‘a’**

**Supplemental Figure 6.**
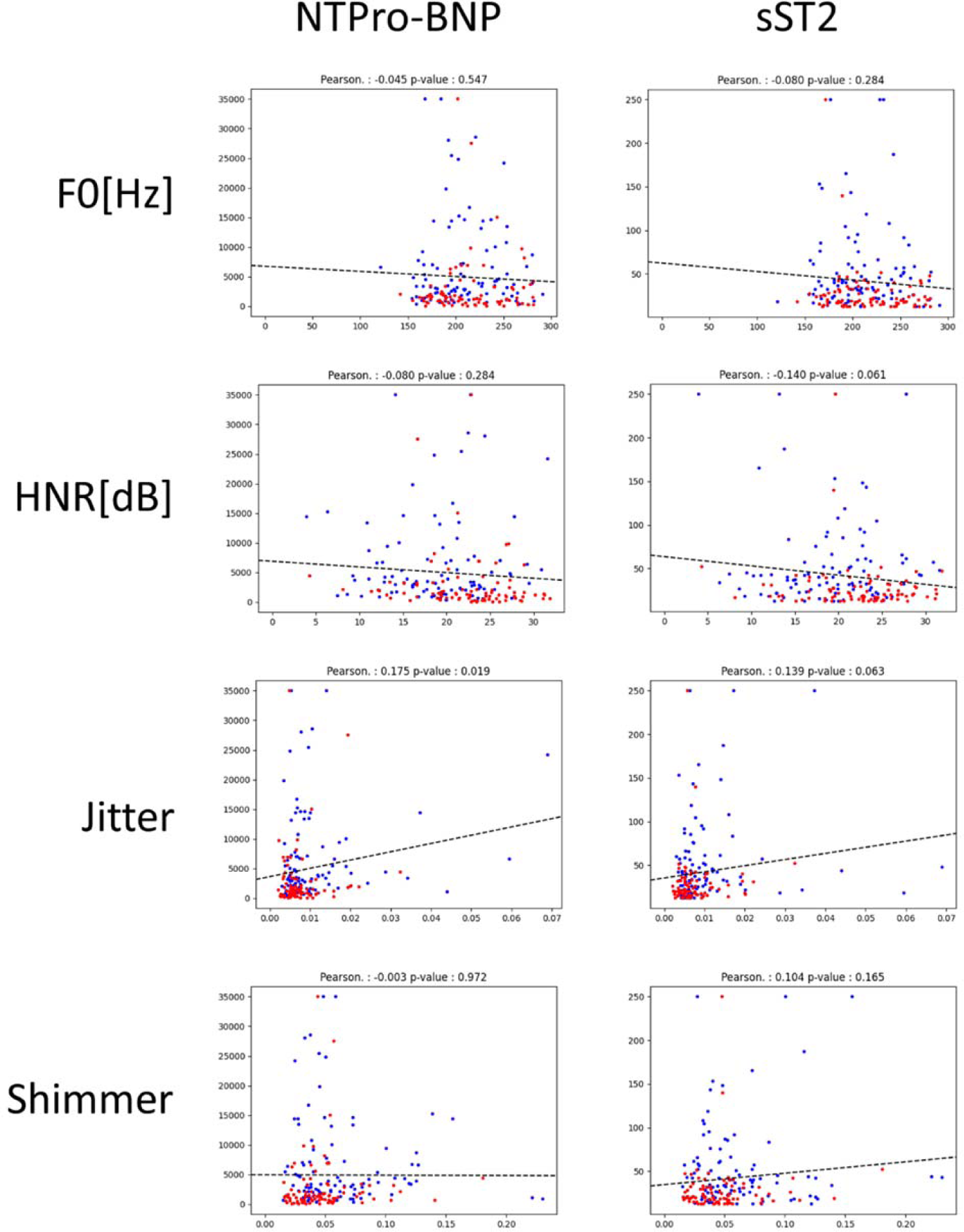
Mel-Spectrogram of the Training Set.

**Supplemental Figure 7.**
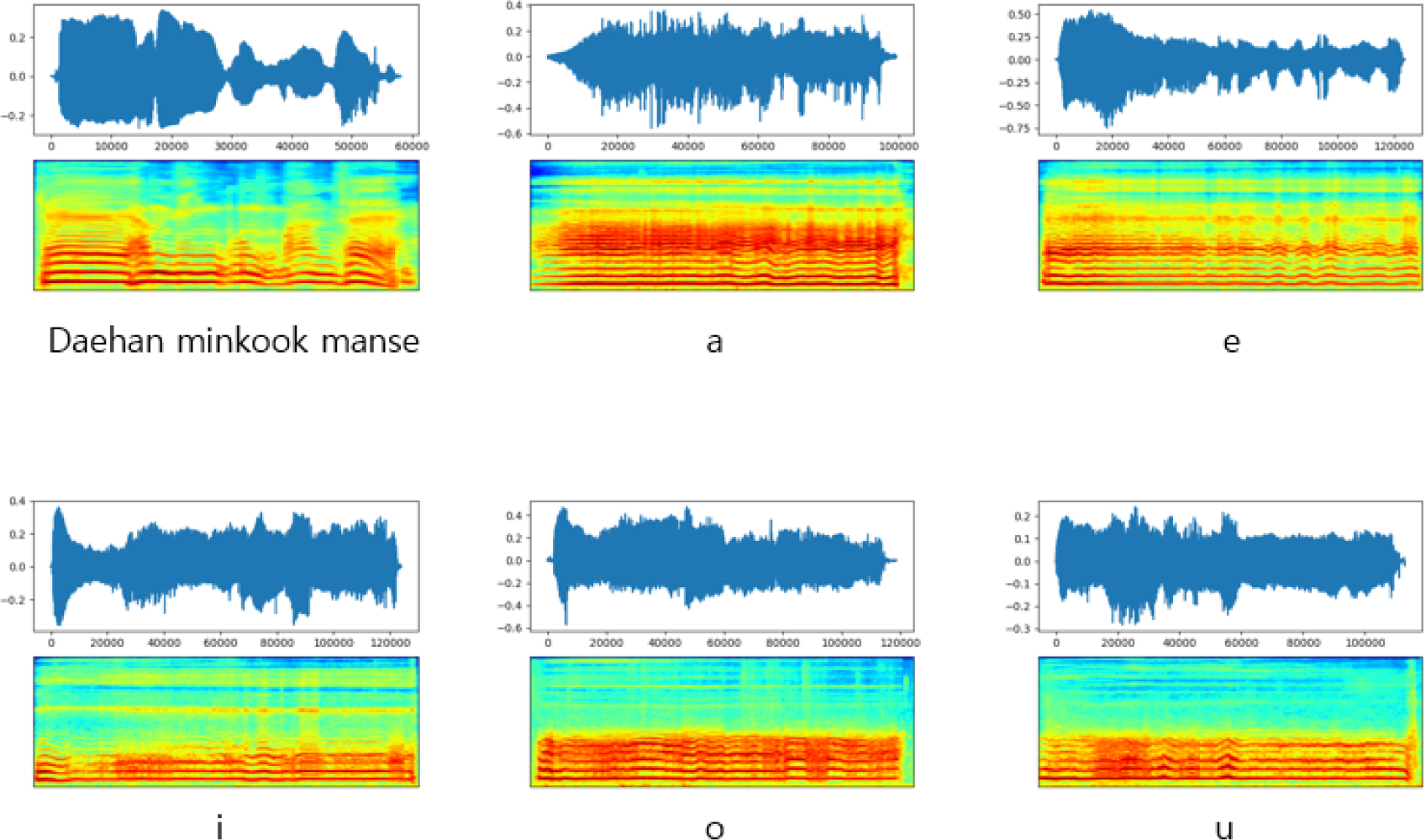
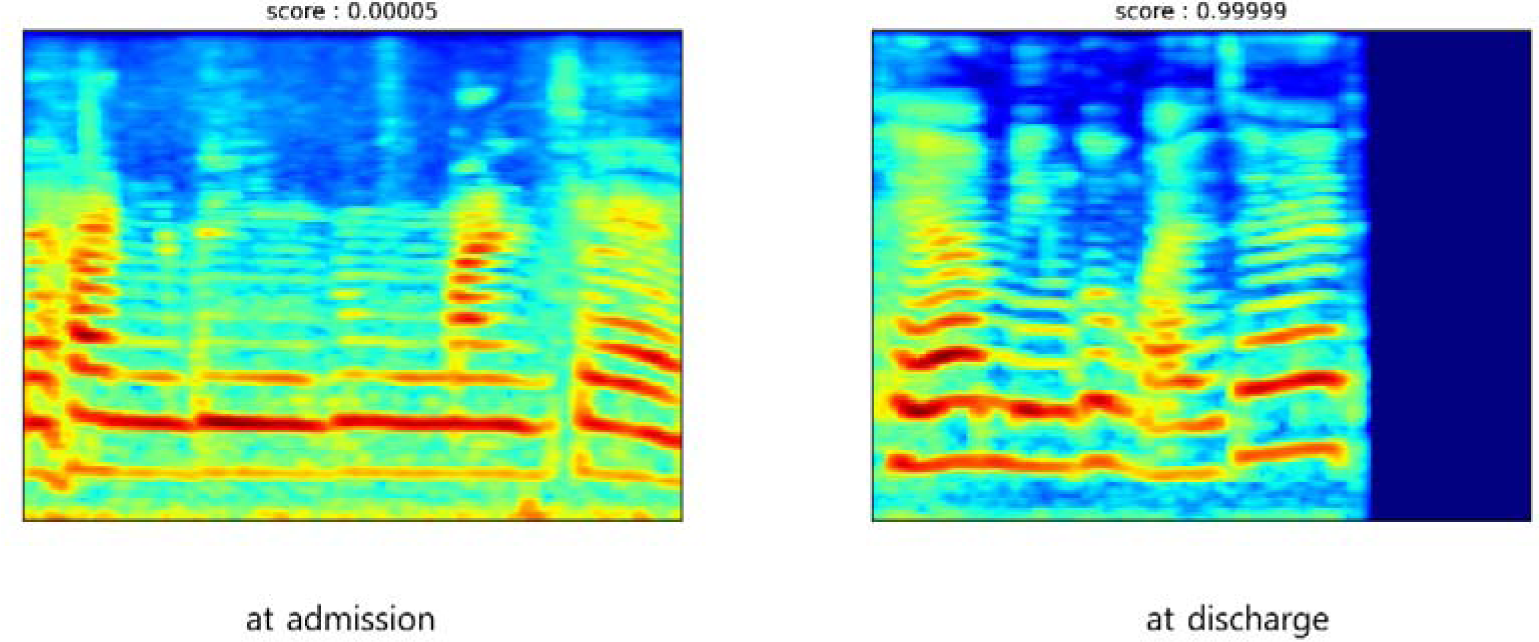
Mel-Spectrograms with the Highest Classification Score and the Lowest Classification Score for ‘dry’ States.

